# SARS-CoV-2 antibody prevalence and determinants of six ethnic groups living in Amsterdam, the Netherlands: a population-based cross-sectional study, June-October 2020

**DOI:** 10.1101/2021.03.08.21252788

**Authors:** Liza Coyer, Anders Boyd, Janke Schinkel, Charles Agyemang, Henrike Galenkamp, Anitra D M Koopman, Tjalling Leenstra, Eric P Moll van Charante, Bert-Jan H van den Born, Anja Lok, Arnoud Verhoeff, Aeilko H Zwinderman, Suzanne Jurriaans, Lonneke A van Vught, Karien Stronks, Maria Prins

## Abstract

**Background:** Ethnic minorities have higher rates of SARS-CoV-2 diagnoses, but little is known about ethnic differences in past exposure. We aimed to determine whether prevalence and determinants of SARS-CoV-2 exposure varied between six ethnic groups in Amsterdam, the Netherlands.

**Methods:** Participants aged 25-79 years enrolled in a population-based prospective cohort were randomly selected within ethnic groups and invited to test for SARS-CoV-2-specific antibodies and answer COVID-19 related questions. We estimated prevalence and determinants of SARS-CoV-2 exposure within ethnic groups using survey-weighted logistic regression adjusting for age, sex and calendar time.

**Results:** Between June 24-October 9, 2020, we included 2497 participants. Adjusted SARS-CoV-2 seroprevalence was comparable between ethnic-Dutch (25/498; 5.5%, 95%CI=3.2-7.9), South-Asian Surinamese (22/451; 4.8%, 95%CI=2.1-7.5), African Surinamese (22/400; 8.2%, 95%CI=3.0-13.4), Turkish (30/408; 7.8%, 95%CI=4.3-11.2) and Moroccan (32/391; 7.0%, 95%CI=4.0-9.9) participants, but higher among Ghanaians (95/327; 26.5%, 95%CI=18.7-34.4). 57.1% of SARS-CoV-2-positive participants did not suspect or were unsure of being infected, which was lowest in African Surinamese (18.2%) and highest in Ghanaians (90.5%). Determinants of SARS-CoV-2 exposure varied across ethnic groups, while the most common determinant was having a household member suspected of infection. In Ghanaians, seropositivity was associated with older age, larger household sizes, living with small children, leaving home to work and attending religious services.

**Conclusions:** No remarkable differences in SARS-CoV-2 seroprevalence were observed between the largest ethnic groups in Amsterdam after the first wave of infections. The higher infection seroprevalence observed among Ghanaians, which passed mostly unnoticed, warrants wider prevention efforts and opportunities for non-symptom-based testing.

## Introduction

Data from the United Kingdom (UK) and United States (US) suggest that certain ethnic minority populations have been disproportionally affected by the coronavirus disease 2019 (COVID-19), caused by SARS-CoV-2. In both countries, a relatively higher number of SARS-CoV-2 polymerase chain reaction (PCR)-positive or clinically-diagnosed COVID-19 cases were observed among ethnic minority groups, particularly people of African and Asian descent.[1-3] The underlying causes for these disparities might include work-related exposure, housing conditions, access to healthcare, help-seeking behavior, and language proficiency.[4-6]

Little is known about ethnic differences in SARS-CoV-2 infections outside the UK and US. This is of particular concern for larger cities in Europe, including the Dutch capital Amsterdam, where half the population comprises migrants, including people with foreign-born parents.[7] Amsterdam witnessed its first confirmed case of SARS-CoV-2 on February 29, 2020 and by December 31, 2020, there were more than 50,000 confirmed infections, 1300 COVID-19-related hospitalizations and 500 COVID-19-related deaths.[8] If SARS-CoV-2 infection prevalence is increased in specific ethnic groups, targeted prevention measures could be instated to help minimize the risk of further transmission.

Ethnic differences in SARS-CoV-2 infection prevalence could be studied using COVID-19 notification registries. However, since the testing policy in the Netherlands has changed several times and until June 1, 2020, testing was largely restricted to symptomatic health care workers or those living or working in long-term care facilities, these data are prone to differential testing uptake. Ethnic differences in testing uptake could be further exacerbated by testing access, willingness to test and disease perceptions. Another limitation of registries is that migration background is often missing. Other data are therefore needed to estimate seroprevalence within specific ethnic groups in Amsterdam.

The Healthy life in an Urban Setting (HELIUS) study is a large, population-based cohort study among six different ethnic groups, which was established with the aim to investigate mechanisms underlying the impact of ethnicity on communicable and non-communicable diseases.[9] From individuals actively enrolled in this study, we determined the prevalence and determinants of exposure to SARS-CoV-2 between the largest ethnic groups in Amsterdam.

## Methods

### Study design and population

The HELIUS study is a multiethnic cohort study conducted in Amsterdam, the Netherlands, which focuses on cardiovascular disease, mental health, and infectious diseases. Detailed procedures have been previously described.[9] Briefly, HELIUS includes persons of Dutch, South-Asian Surinamese, African Surinamese, Ghanaian, Moroccan, and Turkish origin, aged between 18 and 70 years at inclusion. A random sample of persons, stratified by ethnic origin, was taken from the municipality register of Amsterdam and subjects were invited to participate. Between January 2011 and December 2015, a total of 24,789 individuals were included.[9] Participants filled in a self-administered questionnaire and underwent a physical examination during which biological samples were obtained. Ethical approval for the HELIUS study was obtained from the Academic Medical Center Ethical Review Board. All participants provided written informed consent.

Ethnicity was defined according to the country of birth of the participant and their parents.[9] Participants were considered to be of non-Dutch ethnic origin if (i) they were born abroad and had at least one parent born abroad (first generation) or (ii) they were born in Netherlands but both their parents were born abroad (second generation). Participants of Dutch origin were born in the Netherlands with both parents who were born in the Netherlands. Surinamese participants were further classified as African Surinamese, South-Asian Surinamese, and Javanese/other/unknown Surinamese, based on self-reporting.

A cross-sectional, serological substudy was performed in participants of the HELIUS study from 24 June to 9 October 2020. Participants were randomly selected within each ethnic group and asked to participate in the substudy. Serum samples for assessment of SARS-CoV-2 antibodies were collected by venipuncture and stored at −20°C. Trained interviewers asked participants questions on uptake of COVID-19-related prevention measures, potential exposure, infection, symptoms and disease.

### Outcomes

SARS-CoV-2 exposure was determined by the presence of SARS-CoV-2 antibodies. SARS-CoV-2-specific antibodies were determined using the WANTAI SARS-CoV-2 Ab Elisa (Wantai Biological Pharmacy Enterprise Co., Beijing, China) according to the manufacturer’s instructions. This Elisa detects IgA, IgM and IgG against the receptor binding domain of the S-protein of SARS-CoV-2.[10]

### Determinants

We defined the following potential determinants: *from the baseline visit of the HELIUS study*– demographics (i.e. age, sex, ethnicity, migration generation, city district), socio-economic factors (i.e. educational level, working status, occupational level, number of people in household), access-to-healthcare indicators (i.e. proficiency with Dutch language, health literacy); *from the COVID-19 substudy visit*–job setting, household members, suspected being infected, thinking household member/steady partner was infected, household member hospitalized for COVID-19, type of people living in household, travelling abroad in 2020 and COVID-19 behaviors in the past week (i.e. number of times leaving the house, type of locations visited, number of visitors, frequency of using public transportation).

### Statistical analysis

SARS-CoV-2 seroprevalence, along with 95% confidence intervals (CI), was modeled per ethnic group using univariable logistic regression. Seroprevalence was then modeled per ethnic group while correcting for sampling, accounting for the population structure of ethnic groups in Amsterdam (i.e. post-stratification), and adjusting for differences in age, sex and calendar time (before/after 15 August 2020, based on the onset of the second wave of SARS-CoV-2 infections in the Netherlands[8]) between ethnic groups. For sampling, the probability of being invited for the COVID-19 substudy (as the proportion of participants invited among those in active follow-up in the parent study) was calculated, as was the conditional probability of participating in the COVID-19 substudy (given the participant’s ethnicity, age, educational level, working status and health literacy). The product of the two probabilities was taken and the inverse of this result, standardized to one, was used as a sampling weight. For post-stratification, a weight was assigned corresponding to the proportion representing the Amsterdam population of each stratum of age (20-44, 45-54, 55-59, 60-79 years), sex (male, female) and ethnicity (Surinamese, Ghanaian, Moroccan, Turkish, Dutch). Sampling and post-stratification weights were placed in a multivariable logistic regression model with covariates ethnicity, age, sex, and calendar time. Given the weighting scheme of this study, variance was calculated with the designed-based Taylor series linearization method using the ‘svy’ commands in STATA. Differences between ethnic groups were tested in the model using the Wald χ2 test.

Seroprevalence was regressed on age (in restricted cubic splines with 3 knots) with sample and post-stratification weights, within subpopulations of ethnic groups. The mean and 95%CI of predicted seroprevalence was plotted over age in years.

To identify determinants of past SARS-CoV-2 infection within ethnic groups, univariable associations between potential determinants and SARS-CoV-2 seropositivity were evaluated. The odds ratios (OR) comparing the odds of seroprevalence across levels of each determinant, and their 95% confidence intervals (CI), were estimated using logistic regression. P-values were obtained using the Wald χ^2^ test. All covariates with a *P*-value≤0.2 in univariable analyses were then included in a multivariable model and after assessing covariate distributions and collinearity, variables with a *P*-value≥0.05 above this threshold were removed in backwards-stepwise fashion until only variables with a *P*-value<0.05 were retained in a final multivariable model. All models included sampling and post-stratification weights. We forced calendar time in all models.

Statistical significance was defined at a *P*-value<0·05. All analyses were conducted using Stata 15.1 (StataCorp, College Station, TX, USA).

## Results

### Study population

Of the 16,889 HELIUS participants who were in active follow-up in 2019-2020, 11,080 (65.6%) were invited (Figure 1). Of these, 2497 (22.5%) were included in the COVID-19 substudy. The response rate varied across ethnic groups, from 15.3-17.2% among Ghanaian, Turkish or Moroccan participants to 49.9% among Dutch participants. Detailed information on differences between HELIUS participants who were and were not invited, and between invited participants who were and were not included, are presented in Supplementary Table 1. Briefly, invited individuals who were included had obtained a slightly higher educational level, were more likely to be employed and were more likely to have adequate health literacy level compared to those who were invited but not included.

**Figure 1.**
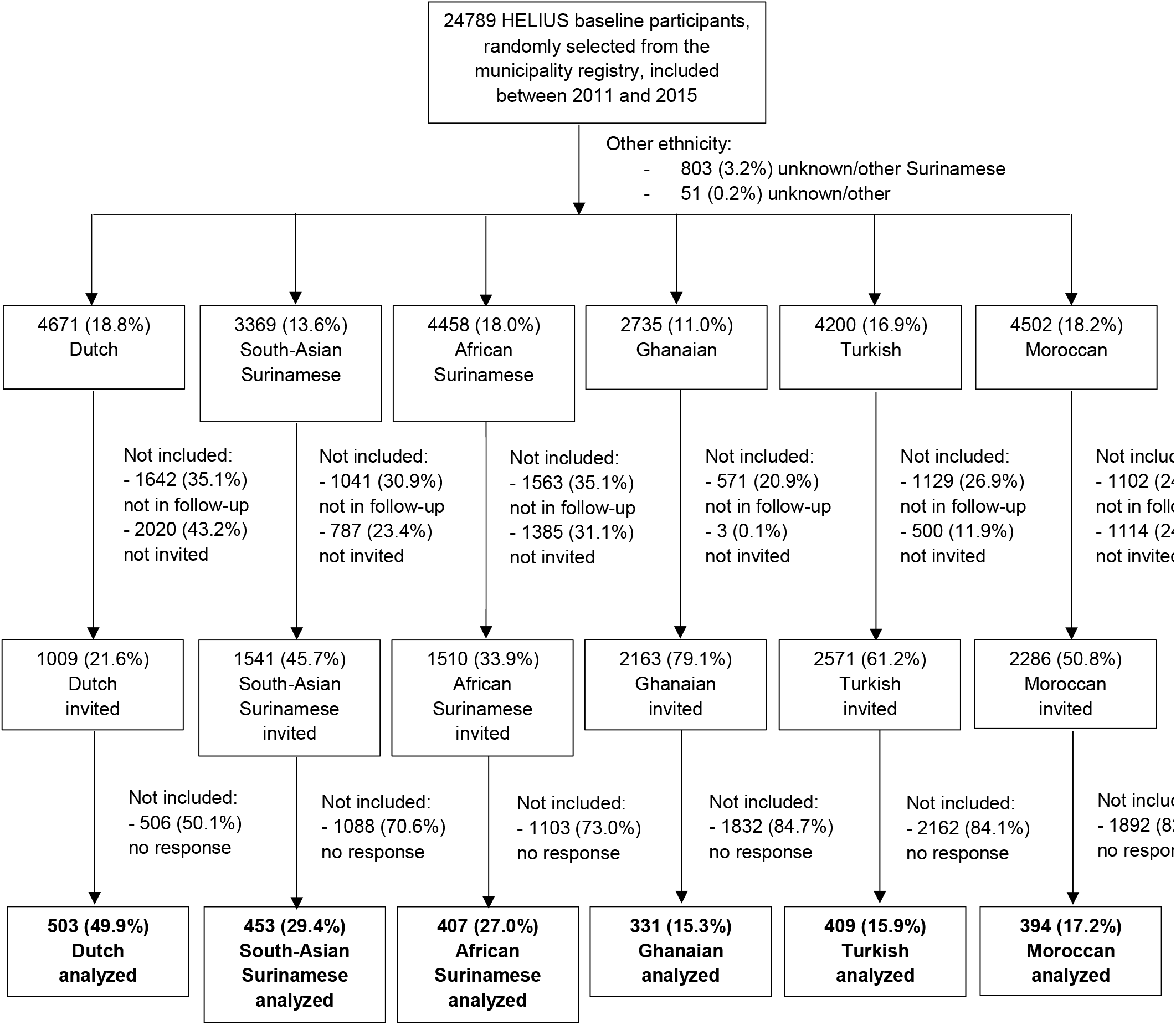
Flowchart depicting the selection of HELIUS participants in the COVID-19 study, Amsterdam, the Netherlands, 24 June - 9 October 2020.

Number included per month within ethnic groups is presented in Supplementary Figure 1. Of 2497 included participants, 503 (20.1%) were of Dutch origin, 453 (18.1%) South-Asian Surinamese, 407 (16.3%) African Surinamese, 331 (13.3%) Ghanaian, 409 (16.4%) Turkish and 394 (15.8%) Moroccan (Supplementary Table 1, Table 1). The median age of included participants was 54 (interquartile range [IQR]: 44-61) and 56.6% were female. In the 1994 participants of non-Dutch origin, the percentage of first-generation migrants was lowest in the Turkish group (74.8%) and highest in the Ghanaian group (98.2%). Dutch participants were the most likely to have a higher vocational or university degree (67.0%) and be employed (75.5%) compared to other ethnicities.

**Table 1.** Characteristics of the HELIUS participants included in the COVID-19 study, by ethnic group (N=2497), Amsterdam, the Netherlands, 24 June - 9 October 2020

| Characteristic | Dutch<br>(n=503) | South-Asian<br>Surinamese<br>(n=453) | African<br>Surinamese<br>(n=407) | Ghanaian<br>(n=331) | Turkish<br>(n=409) | Moroccan<br>(n=392) | P-value |
| --- | --- | --- | --- | --- | --- | --- | --- |
|  | n (%) | n (%) | n (%) | n (%) | n (%) | n (%) |  |
| **Sex** |  |  |  |  |  |  |  |
| Male | 237 (47.1%) | 179 (39.5%) | 165 (40.5%) | 145 (43.8%) | 184 (45.0%) | 173 (43.9%) | 0.19 |
| Female | 266 (52.9%) | 274 (60.5%) | 242 (59.5%) | 186 (56.2%) | 225 (55.0%) | 221 (56.1%) |  |
| **Age in years on 1 January 2020** |  |  |  |  |  |  |  |
| Median [IQR] | 57 [45-66] | 56[47-63] | 59 [50-65] | 54 [47-59] | 48 [40-56] | 49 [39-56] | <0.001 |
| **Migration generation** |  |  |  |  |  |  |  |
| 1<sup>st</sup> | N.A. | 370 (81.7%) | 355 (87.2%) | 325 (98.2%) | 306 (74.8%) | 300 (76.1%) | <0.001 |
| 2<sup>nd</sup> | N.A. | 83 (18.3%) | 52 (12.8%) | 6 (1.8%) | 103 (25.2%) | 94 (23.9%) |  |
| **City district<sup>a</sup>** |  |  |  |  |  |  |  |
| Centre | 88 (17.5%) | 18 (4.0%) | 15 (3.7%) | 5 (1.5%) | 4 (1.0%) | 12 (3.0%) | <0.001 |
| East | 99 (19.7%) | 53 (11.7%) | 84 (20.6%) | 25 (7.6%) | 66 (16.1%) | 94 (23.9%) |  |
| West | 89 (17.7%) | 6 (1.3%) | 34 (8.4%) | 19 (5.7%) | 66 (16.1%) | 83 (21.1%) |  |
| South | 112 (22.3%) | 32 (7.1%) | 28 (6.9%) | 8 (2.4%) | 30 (7.3%) | 38 (9.6%) |  |
| New-West | 45 (8.9%) | 111 (24.5%) | 51 (12.5%) | 18 (5.4%) | 231 (56.5%) | 147 (37.3%) |  |
| Southeast | 65 (12.9%) | 229 (50.6%) | 192 (47.2%) | 254 (76.7%) | 6 (1.5%) | 19 (4.8%) |  |
| Other/missing | 5 (1.0%) | 4 (0.9%) | 3 (0.7%) | 2 (0.6%) | 6 (1.5%) | 1 (0.3%) |  |
| **Educational level<sup>a</sup>** |  |  |  |  |  |  |  |
| No school/elementary school | 10 (2.0%) | 56 (12.4%) | 15 (3.7%) | 78 (23.6%) | 78 (19.1%) | 90 (22.8%) | <0.001 |
| Lower vocational/lower<br>secondary school | 56 (11.1%) | 156 (34.4%) | 124 (30.5%) | 128 (38.7%) | 84 (20.5%) | 64 (16.2%) |  |
| Intermediary<br>vocational/intermediary<br>secondary school | 99 (19.7%) | 137 (30.2%) | 142 (34.9%) | 73 (22.1%) | 124 (30.3%) | 125 (31.7%) |  |
| Higher vocational/university | 337 (67.0%) | 103 (22.7%) | 124 (30.5%) | 26 (7.9%) | 108 (26.4%) | 94 (23.9%) |  |
| Missing | 1 (0.2%) | 1 (0.2%) | 2 (0.5%) | 26 (7.9%) | 15 (3.7%) | 21 (5.3%) |  |
| **Labor participation<sup>b</sup>** |  |  |  |  |  |  |  |
| Employed | 380 (75.5%) | 308 (68.0%) | 292 (71.7%) | 203 (61.3%) | 247 (60.4%) | 229 (58.1%) | <0.001 |
| Not in workforce | 90 (17.9%) | 47 (10.4%) | 40 (9.8%) | 10 (3.0%) | 59 (14.4%) | 63 (16.0%) |  |
| Unemployed/on benefits | 21 (4.2%) | 53 (11.7%) | 47 (11.5%) | 60 (18.1%) | 62 (15.2%) | 57 (14.5%) |  |
| Disabled | 11 (2.2%) | 39 (8.6%) | 24 (5.9%) | 28 (8.5%) | 27 (6.6%) | 22 (5.6%) |  |
| Unknown/missing | 1 (0.2%) | 6 (1.3%) | 4 (1.0%) | 30 (9.0%) | 14 (3.4%) | 23 (5.8%) |  |
| **Occupational level<sup>a</sup>** |  |  |  |  |  |  |  |
| Elementary occupations | 5 (1.0%) | 36 (7.9%) | 22 (5.4%) | 162 (48.9%) | 52 (12.7%) | 46 (11.7%) | <0.001 |
| Lower occupations | 46 (9.1%) | 127 (28.0%) | 101 (24.8%) | 69 (20.8%) | 102 (24.9%) | 92 (23.4%) |  |
| Intermediary occupations | 107 (21.3%) | 143 (31.6%) | 146 (35.9%) | 21 (6.3%) | 88 (21.5%) | 94 (23.9%) |  |
| Higher occupations | 203 (40.4%) | 79 (17.4%) | 91 (22.4%) | 11 (3.3%) | 51 (12.5%) | 65 (16.5%) |  |
| Scientific occupations | 115 (22.9%) | 20 (4.4%) | 19 (4.7%) | 6 (1.8%) | 32 (7.8%) | 10 (2.5%) |  |
| Missing | 27 (5.4%) | 48 (10.6%) | 28 (6.9%) | 62 (18.7%) | 84 (22.5%) | 87 (22.1%) |  |
| **Job setting<sup>b</sup>** |  |  |  |  |  |  |  |
| No job / caretaker only | 117 (23.3%) | 144 (31.8%) | 120 (29.5%) | 90 (27.2%) | 138 (33.7%) | 132 (33.5%) | <0.001 |
| Job with no contact within 1.5 meter | 96 (19.1%) | 65 (14.3%) | 39 (9.6%) | 66 (19.9%) | 67 (16.4%) | 54 (13.7%) |  |
| Other job with contact within 1.5 meter | 145 (28.8%) | 154 (34.0%) | 131 (32.2%) | 115 (34.7%) | 130 (31.8%) | 114 (28.9%) |  |
| Child care/schools/higher education | 62 (12.3%) | 27 (6.0%) | 43 (10.6%) | 10 (3.0%) | 25 (6.1%) | 48 (12.2%) |  |
| Bar/restaurant | 12 (2.4%) | 10 (2.2%) | 11 (2.7%) | 23 (6.9%) | 6 (1.5%) | 7 (1.8%) |  |
| Hospital/long-term care facility/Health care worker elsewhere | 71 (14.1%) | 51 (11.3%) | 63 (15.5%) | 26 (7.9%) | 41 (10.0%) | 36 (9.1%) |  |
| Missing | 0 (0.0%) | 2 (0.4%) | 0 (0.0%) | 1 (0.3%) | 2 (0.5%) | 3 (0.8%) |  |
| **Difficulty with Dutch language<sup>a</sup>** |  |  |  |  |  |  |  |
| No | N.A. | 348 (76.8%) | 359 (88.2%) | 41 (12.4%) | 189 (46.2%) | 211 (53.6%) | <0.001 |
| Yes | N.A. | 104 (23.0%) | 46 (11.3%) | 264 (79.8%) | 206 (50.4%) | 162 (41.1%) |  |
| Missing | N.A. | 1 (0.2%) | 2 (0.5%) | 26 (7.9%) | 14 (3.4%) | 21 (5.3%) |  |
| **Health literacy (SBSQ)<sup>a</sup>** |  |  |  |  |  |  |  |
| Adequate | 500 (99.4%) | 437 (96.5%) | 400 (98.3%) | 209 (63.1%) | 310 (75.8%) | 308 (78.2%) | <0.001 |
| Low | 3 (0.6%) | 16 (3.5%) | 7 (1.7%) | 97 (29.3%) | 87 (21.3%) | 64 (16.2%) |  |
| Missing | 0 (0.0%) | 0 (0.0%) | 0 (0.0%) | 25 (7.6%) | 12 (2.9%) | 22 (5.6%) |  |
| **Diabetes mellitus<sup>c</sup>** |  |  |  |  |  |  |  |
| No | 478 (95.0%) | 358 (79.0%) | 362 (88.9%) | 297 (89.7%) | 366 (89.5%) | 345 (87.6%) | <0.001 |
| Yes | 18 (3.6%) | 88 (19.4%) | 42 (10.3%) | 30 (9.1%) | 35 (8.6%) | 41 (10.4%) |  |
| Missing | 7 (1.4%) | 7 (1.5%) | 3 (0.7%) | 4 (1.2%) | 8 (2.0%) | 8 (2.0%) |  |
| **High blood pressure<sup>d</sup>** |  |  |  |  |  |  |  |
| No | 370 (73.6%) | 261 (57.6%) | 198 (48.6%) | 143 (43.2%) | 321 (78.5%) | 305 (77.4%) | <0.001 |
| Yes | 127 (25.2%) | 185 (40.8%) | 207 (50.9%) | 181 (54.7%) | 82 (20.0%) | 81 (20.6%) |  |
| Missing | 6 (1.2%) | 7 (1.5%) | 2 (0.5%) | 7 (2.1%) | 6 (1.5%) | 8 (2.0%) |  |
| **Body Mass Index (kg/m2), median (IQR)<sup>a</sup>** | 24 [22-27] | 25 [23-28] | 27 [24-29] | 28 [25-31] | 27 [24-31] | 27 [24-30] | <0.001 |
| **Month of study visit<sup>b</sup>** |  |  |  |  |  |  |  |
| June | 86 (17.1%) | 44 (9.7%) | 31 (7.6%) | 0 (0.0%) | 0 (0.0%) | 32 (8.1%) | <0.001 |
| July | 265 (52.7%) | 233 (51.4%) | 203 (49.9%) | 38 (11.5%) | 151 (36.9%) | 170 (43.1%) |  |
| August | 108 (21.5%) | 98 (21.6%) | 110 (27.0%) | 135 (40.8%) | 88 (21.5%) | 85 (21.6%) |  |
| September | 39 (7.8%) | 75 (16.6%) | 56 (13.8%) | 125 (37.8%) | 127 (31.1%) | 74 (18.8%) |  |
| October | 5 (1.0%) | 3 (0.7%) | 7 (1.7%) | 33 (10.0%) | 43 (10.5%) | 33 (8.4%) |  |

**Abbreviations:** BMI, body mass index; HELIUS, Healthy Life in an Urban Setting; IQR, interquartile range; N.A., not applicable; SBSQ, Set of Brief Screening Question <sup>a</sup> Presumed higher
exposure categories had priority, i.e. if someone was working in a school and as a health care worker, they were categorized as a health care worker. Caretakers were not included as a
category because many had other jobs.
<sup>a</sup> Measured at baseline (2011-2015) <sup>b</sup> Measured at COVID-1 visit (2020) <sup>c</sup> Based on self-report, increased fasting glucose ( $\geq 7$ mmol/l) or use of glucose-lowering medication <sup>d</sup> Based on self-
report, SBP $\geq 140$ mmHg, DBP $\geq 90$ or blood pressure-lowering medication

### SARS-CoV-2 seroprevalence

Of 2497 included, 2483 (99.4%) participants had a SARS-CoV-2 antibody test result. Of these 2483, 226 were positive, 2249 negative and 8 had an equivocal test result. The distribution of signal-to-cutoff ratios for positive test results is shown per ethnic group in Supplementary Figure 2. The proportion with a positive result did not increase over time in any of the ethnic groups, except for the South-Asian Surinamese group (Supplementary Figure 1).

Unadjusted and adjusted seroprevalence estimates per ethnic group are provided in Figure 2 and Supplementary Table 2. Adjusted seroprevalence was comparable between the Dutch (5.5%, 95%CI=3.2-7.9), South-Asian Surinamese (4.8%, 95%CI=2.1-7.5), African Surinamese (8.2%, 95%CI=3.0-13.4), Turkish (7.8%, 95%CI=4.3-11.2) and Moroccan (7.0%, 95%CI=4.0-9.9) groups, but higher in the Ghanaian group compared to all other groups (26.5%, 95%CI=18.7-34.4, *P*<0.001).

**Figure 2.**
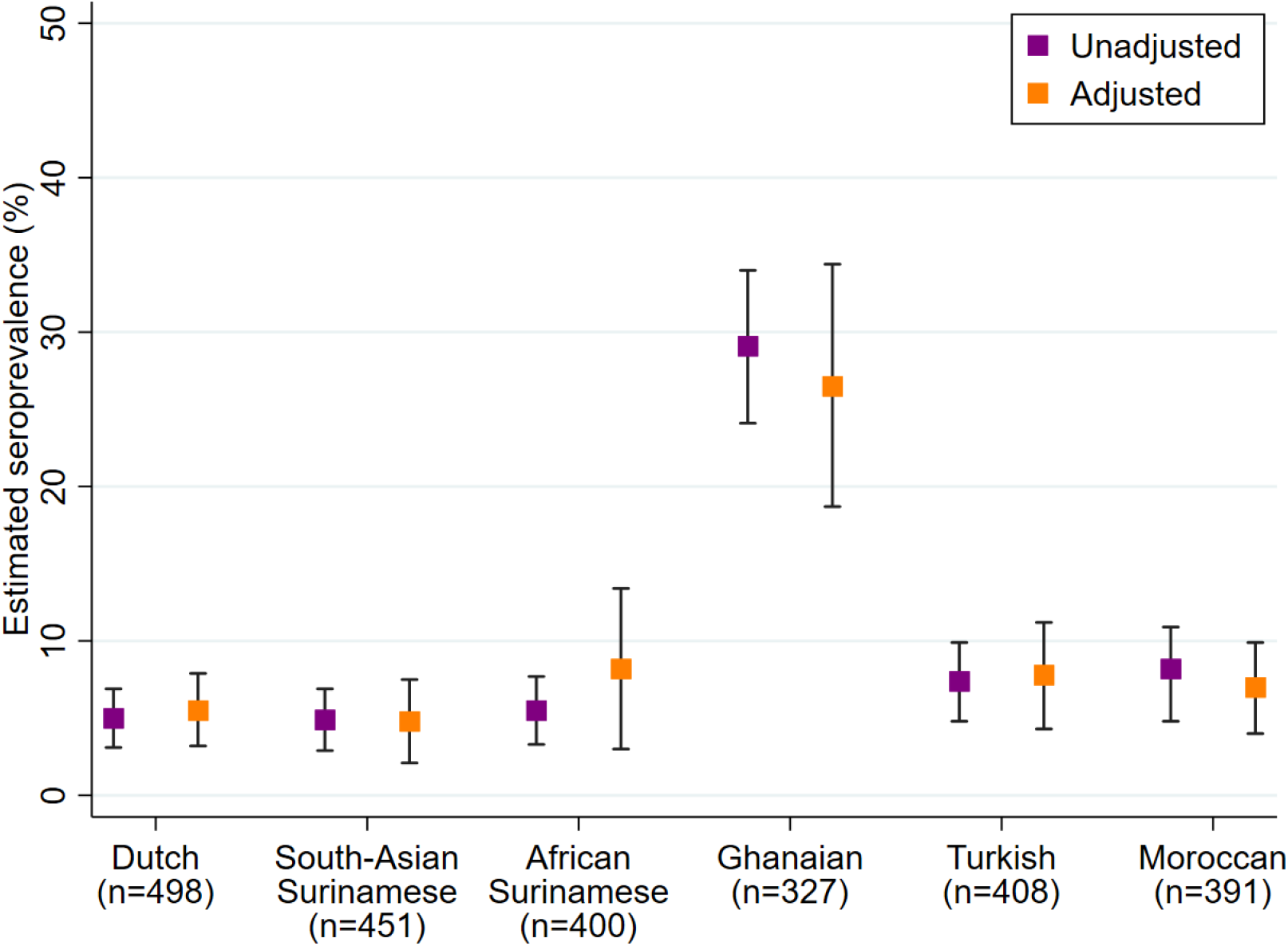
Unadjusted and adjusted SARS-CoV-2 seroprevalence per ethnic group (N=2475), Amsterdam, the Netherlands, 24 June - 9 October 2020. Footnote: We excluded individuals with an equivocal result (n=8) from the seroprevalence calculation. Boxes represent the seroprevalence estimate, bands the corresponding 95% confidence interval. Adjusted seroprevalence estimates were corrected for sampling, accounted for the population structure of ethnic groups in Amsterdam (i.e. post-stratification), and adjusted for differences in age, sex and calendar time (before/after 15 August 2020) between ethnic groups.

Figure 3 shows adjusted seroprevalence estimates as a function of age in years for each ethnic group. In the African Surinamese group, seroprevalence decreased with age. In the Ghanaian group, the highest seroprevalence was observed between the ages of 50-55 years.

**Figure 3.**
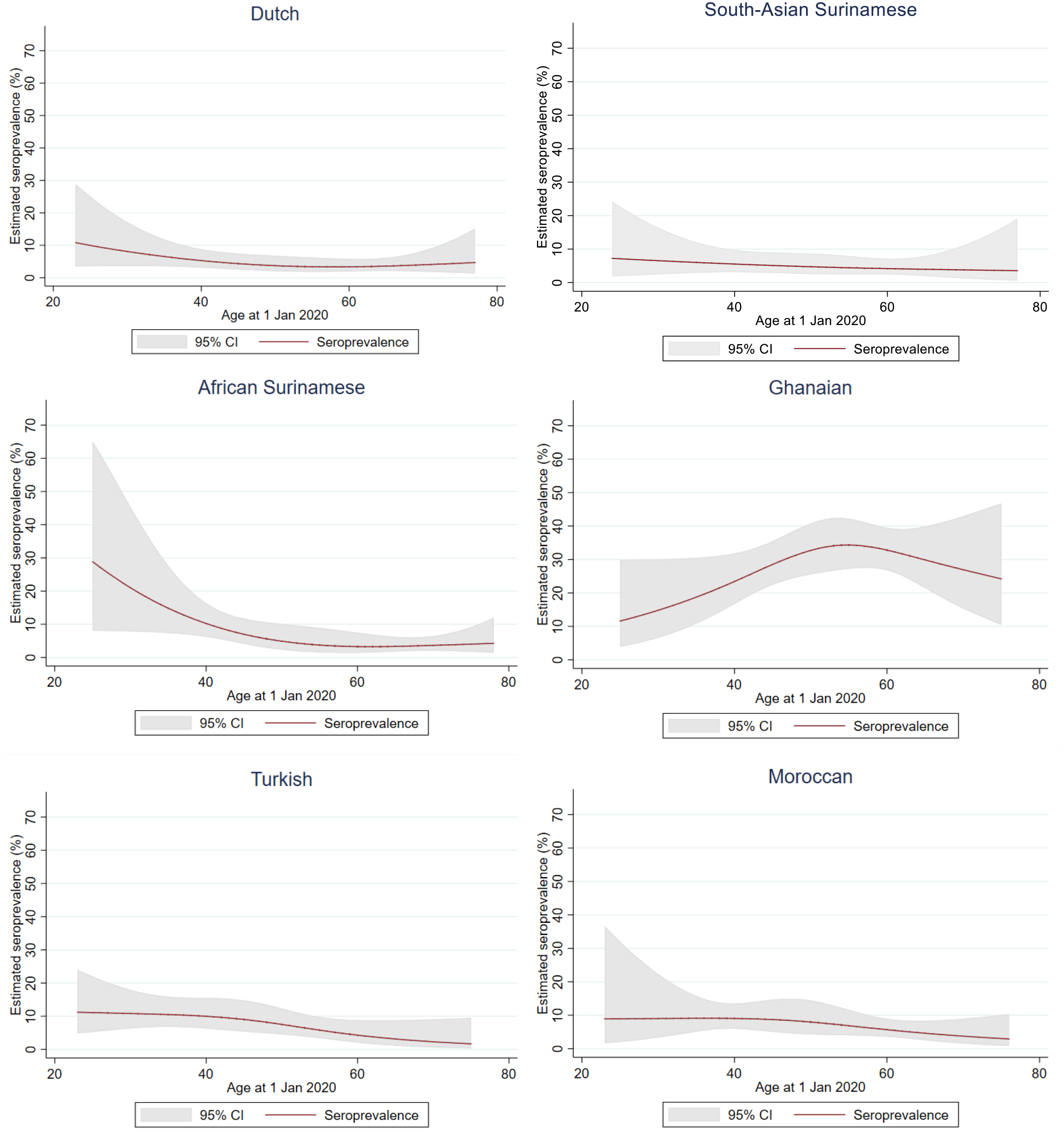
SARS-CoV-2 seroprevalence and age by ethnic group, Amsterdam, the Netherlands, 24 June - 9 October 2020. Footnote: Seroprevalence was regressed on age (in restricted cubic splines with 3 knots) with sample and post-stratification weights, within subpopulations of ethnic groups.

### COVID-19-related symptoms

Table 2 describes SARS-CoV-2-related characteristics of included participants. Of 2497 participants, 348 (13.9%) suspected being infected with SARS-CoV-2, 2144 (85.9%) did not suspect or were unsure of being infected. 90.5% of Ghanaian participants who tested positive did not suspect or were unsure of being infected, mainly because most of these individuals had not experienced symptoms (58.7%). SARS-CoV-2 positive individuals from other ethnic groups more frequently suspected being infected (range 59.1% to 81.8%).

**Table 2.**
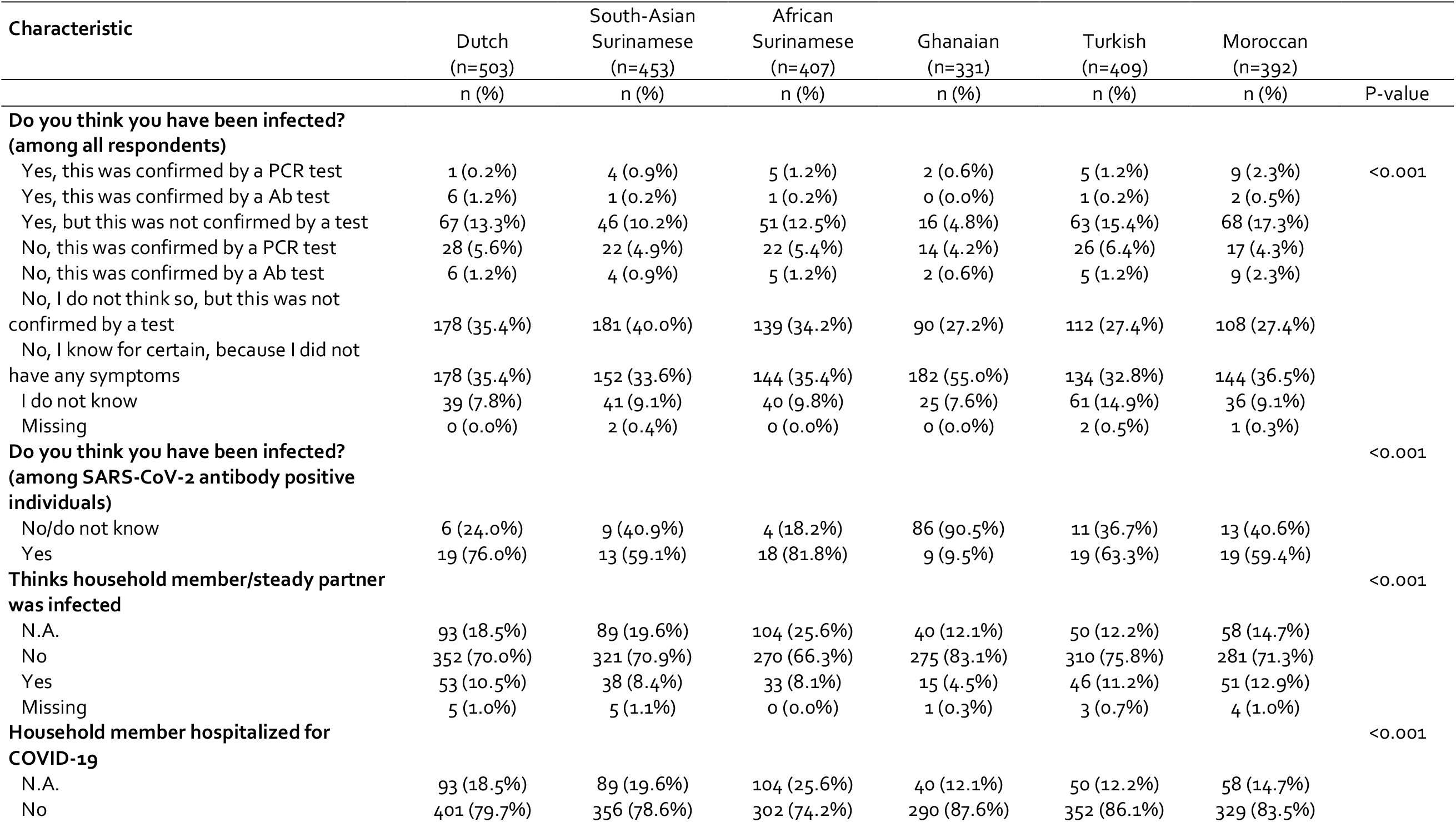
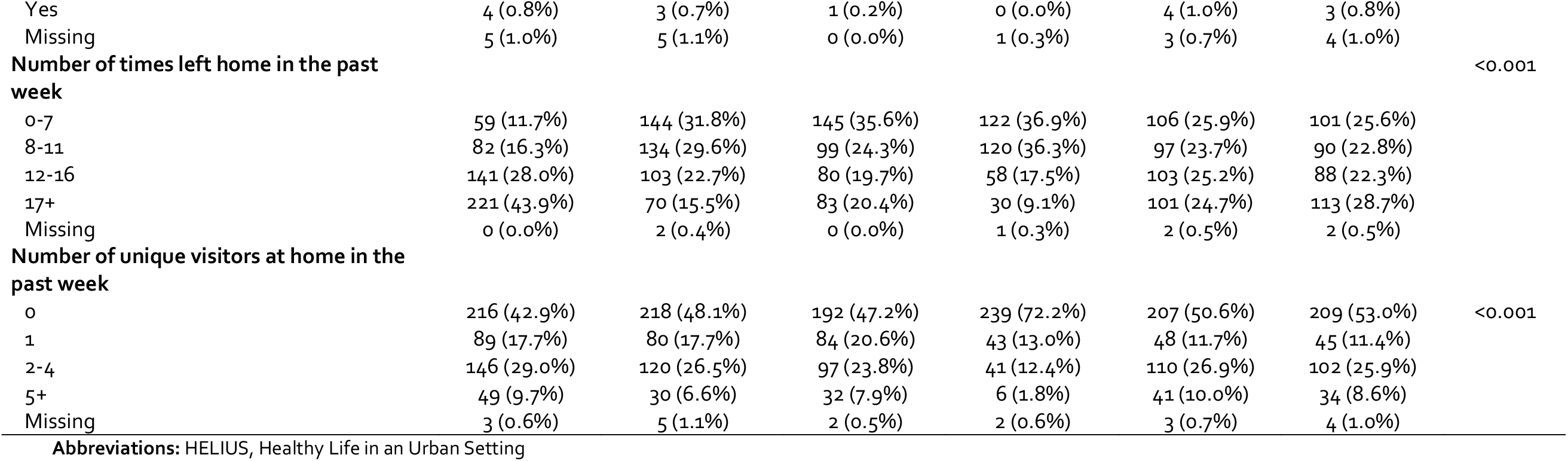
SARS-CoV-2-related characteristics of the HELIUS participants included in the COVID-19 study, by ethnicity (N=2497), Amsterdam, the Netherlands, 24 June - 9 October 2020

### Determinants of SARS-CoV-2 seropositivity per ethnic group

Univariable analysis of determinants of SARS-CoV-2 seropositivity is presented per ethnic group in Supplementary Tables 2-7. In multivariable analysis (Figure 4), having a household member suspected of infection was associated with SARS-CoV-2 seropositivity in Dutch, South-Asian Surinamese, Turkish and Moroccan participants. Recently traveling abroad was associated with seropositivity in Dutch and South-Asian Surinamese participants. In Ghanaian participants, older age, increasing household size, living with children ≤3 years old, and leaving home to work and attending religious services were associated with SARS-CoV-2 seropositivity. Increased odds for SARS-CoV-2 seropositivity were also observed for leaving home to pick up medication or visiting a doctor in the past week (Dutch participants), living with other adults (African Surinamese), having had ≥2 unique visitors in the past week (African Surinamese), leaving home to walk or exercise outside and using public transportation in the past week (Turkish participants) and occupational level (Moroccan participants).

**Figure 4.**
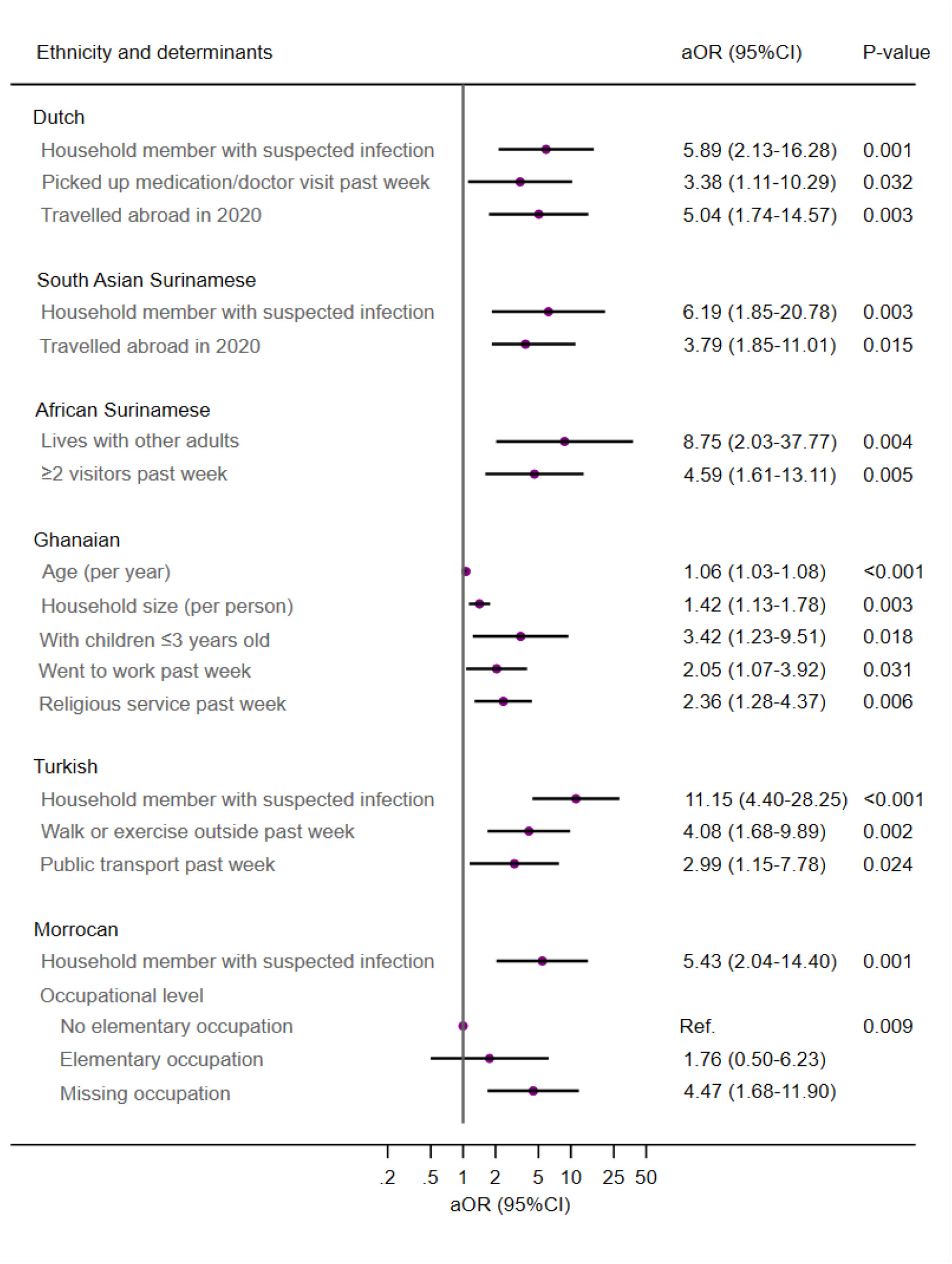
Determinants of SARS-CoV-2 seropositivity by ethnic group, HELIUS COVID-19 study, 24 June - 9 October 2020 (multivariable analysis)

## Discussion

After the first wave of the SARS-CoV-2 epidemic, we observed no evidence of ethnic disparities in past SARS-CoV-2 infection between the six largest ethnic groups residing in Amsterdam, The Netherlands, with the noteworthy exception of individuals of Ghanaian origin. We estimated that 26% of the adult Ghanaian group had developed SARS-COV-2 antibodies, compared to 5-8% of the other adult ethnic groups. Increased risk of past infection was present among individuals who reported a household member suspected of infection in four of the six groups. Amongst other factors, leaving home to work and attending religious services were associated with seropositivity in Ghanaian individuals, while using public transportation was associated with seropositivity in Turkish individuals. Determinants differed between ethnicities, hence demonstrating that broad generalizations of some SARS-CoV-2-related determinants might not be appropriate for individual ethnic groups.

Among the determinants of SARS-CoV-2 seropositivity, work and travelling to work, most likely via public transportation, represents a common theme in individuals of non-Dutch origin. Working from home was one of the first preventive measures introduced in the Netherlands to mitigate spread of SARS-CoV-2.[11] However, this was not feasible for individuals with lower professional levels and jobs requiring physical presence, many of whom were of non-Dutch origin. Interestingly, Moroccan individuals in the missing occupation category appeared to be more often seropositive. Previous research suggests that the health of individuals in this category resembles that of individuals with elementary or intermediary professions,[12] implying that working conditions could put these individuals at risk of infection.

Although attending religious services was asked only for the past week and infections may have occurred as early as in March 2020, exposure to SARS-CoV-2 during attendance at religious services might have driven many of the past infections observed in the Ghanaian group. Religious services, along with demonstrations, were allowed to continue without a maximum number of attendees, as stipulated by Dutch law,[13] which could have fostered further spread of SARS-CoV-2. Many places of worship did, however, implement social distancing measures. A nationwide study demonstrated similar findings in that Orthodox-Reformed Protestants were at increased risk for SARS-CoV-2 seropositivity during the first wave of the pandemic.[14] Increased infection risk for people attending religious services has also been demonstrated in studies from other countries.[15-17]

Strikingly, 90% of Ghanaians with SARS-CoV-2 antibodies did not suspect or were unsure of being infected, many because they did not report experiencing any COVID-19-related symptoms. This is in stark contrast to other ethnic groups in which most SARS-CoV-2 positive individuals had suspected of being infected. If these infections were indeed asymptomatic in Ghanaians, many could have been completely unaware of their infection and as a result, might have carried out their normal routines despite unknowingly continuing transmission. The dense clustering of Ghanaians in the South-East city district of Amsterdam might have also accelerated transmission, as we unknowingly may have sampled a cluster of infections within a specific neighbourhood or religious center. Nevertheless, there were no infection clusters within Ghanaian individuals identified during the first wave by the local Public Health Service (personal communication T. Leenstra, January 27, 2021), when SARS-CoV-2 PCR testing was restricted. Our study clearly indicates that to reduce ongoing and unnoticed transmission of SARS-CoV-2, expanded testing needs to include those groups in which the proportion of asymptomatic individuals might be high, such as the Ghanaian residents of Amsterdam.

Since data from Ghana on SARS-CoV-2 seroprevalence and proportion of asymptomatic infection are limited, we cannot make any distinction on whether our finding reflects the epidemiology in the country of origin or is specific to Ghanaian individuals in the Netherlands. One modelling study suggests that Ghana is one of the four most affected African countries in terms of cases, but has a relatively low death rate.[18] A study among Kenyan blood donors found a surprisingly high seroprevalence (4.3%) from what can be inferred by the low number of COVID-19-related hospitalisations and deaths.[19] Further research is needed to clarify the role of symptom burden, earlier exposure to coronaviruses, or differences in genetic vulnerability to symptoms in explaining the seemingly high proportion of asymptomatic cases in Ghanaians.[20,21]

Having a household member suspected of being infected was the most common and consistent determinant of seropositivity. This finding supports observations that during periods of more extensive lock-downs, most transmissions occur in household settings and is related to symptomatic infection, age distribution and social interactions within households.[22-24] Other household determinants of seropositivity were observed in specific ethnic groups and included living with other adults, living with children ≤3 years old, and larger household sizes.

In the Netherlands, a series of restrictions was introduced in mid-March, when the spread of SARS-CoV-2 was still limited.[11] The finding that seroprevalence did not differ between ethnic groups, other than Ghanaian, implies that these restrictive measures were able to prevent the spread of infection equally across ethnicities. Furthermore, additional data from individuals participating in the parent HELIUS study showed that non-ethnic Dutch groups in general were as likely as ethnic-Dutch to adhere to prevention measures (personal communication F. Chilunga, January, 27 2021). It should be mentioned that our results also stem from a setting where economic inequalities are not prohibitive to healthcare access.[25]

In comparison to the seroprevalence estimates, people from large ethnic groups (Netherlands Antilles, Morocco, Surinam, Turkey, Ghana) had increased hospitalisation rates compared to ethnic Dutch individuals living in Amsterdam between February and May 2020,[26] as shown in other settings.[2,3] In addition, individuals with a migration background living in the Netherlands had a higher excess mortality during the first six weeks of the COVID-19 pandemic.[27] Our data suggest that, apart from Ghanaians, the increased rates of hospitalisations and deaths in non-Dutch ethnic groups during this period cannot be explained by a higher infection rate. The severity of COVID-19 can be impacted to a large extent by underlying comorbidities,[28] which vary across ethnic groups[9] and could explain differences in severity.[29] Healthcare inequalities, racism, stigmatisation and discrimination witnessed by ethnic minorities and differences in healthcare seeking-behaviour may provide additional explanations for these disparities.[30-34]

Strengths of our study include population-based sampling, with a large number of participants from the major ethnic groups living in Amsterdam, representing various levels of socioeconomic status; measuring seroprevalence via antibodies in individuals with and without previous COVID-19-related symptoms; and obtaining individual-level determinants of infection. Nonetheless, there are several limitations. First, our study includes a random subsample of HELIUS participants and there may be selection bias. Undocumented people and other ethnic groups living in Amsterdam were not included in the parent study. Second, participants in our substudy may have been more concerned about their health compared to non-participants. Notwithstanding the differential response rate between ethnicities in this substudy, the distribution of characteristics was largely similar between included and non-included HELIUS participants. Our estimates, corrected for sampling and post-stratification, were also close to those from a nationwide study that included mainly people of Dutch origin and revealed a 6% seroprevalence among the Amsterdam population in June 2020.[35] Data were also collected over a span of 4 months, which reflects different points of the epidemic, and thus the timing of testing could bias estimates. We attempted to mitigate this issue by adjusting for calendar time. Furthermore, prevention measures remained mostly the same and nationwide incidence was quite stable during this period, thereby limiting the effect of this bias.[8,36] Third, as this study was cross-sectional and infection occurred in the past, it is difficult to make any causal inference with respect to determinants. Fourth, fear of stigmatization or consequences for work might have led to an underreporting of suspected past infection and symptoms, particularly among Ghanaians. Finally, circulating SARS-CoV-2 antibodies could have disappeared after infection,[37,38] although this was probably limited during the study period,[39,40] and individuals could not participate in this substudy if they were experiencing COVID-19-related symptoms, both of which likely led to underestimated seroprevalence.

In conclusion, most ethnic groups displayed comparable seroprevalence after the first SARS-CoV-2 wave in Amsterdam, yet the substantially higher prevalence among the smaller Ghanaian population, possibly infections without symptoms, is of concern. Targeted prevention campaigns addressing the needs of specific ethnic groups and expanding testing opportunities are urgently warranted. In addition, prevention measures for those who cannot work from home should be intensified, also by bringing to light the employer’s role in reducing COVID-19 transmissions.

## Data Availability

The HELIUS data are owned by the Amsterdam UMC, location AMC, in Amsterdam, The Netherlands. Any researcher can request the data by submitting a proposal to the HELIUS Executive Board as outlined at http://www.heliusstudy.nl/en/researchers/collaboration, by. The HELIUS Executive Board will check proposals for compatibility with the general objectives, ethical approvals and informed consent forms of the HELIUS study. There are no other restrictions to obtaining the data and all data requests will be processed in the same manner.

## Author contributions

MP, KS, JS and CA conceived, designed or oversaw the study. HG, AK and JS were involved in the acquisition of data. LC and AB conducted the statistical analysis. LC, AB and MP drafted the manuscript. All authors read and approved the final manuscript.

## Declaration of interests

The authors declare that they have no competing interests related to the project.

## Funding

This work was supported by ZonMw (10430022010002) and the Public Health Service of Amsterdam. The HELIUS study is conducted by Amsterdam UMC, location Academic Medical Center and the Public Health Service of Amsterdam. Both organizations provided core support for HELIUS. The HELIUS study is also funded by the Dutch Heart Foundation (2010 T084), ZonMw (200500003), the European Union (FP-7) (278901), and the European Fund for the Integration of non-EU immigrants (EIF) (2013EIF013).

## Acknowledgements

The authors would like to acknowledge the HELIUS COVID-19 study participants for their contribution and the HELIUS team for data collection and management. They would also like to thank Anton Janssen for providing population tables.

## Notes

### Competing Interest Statement

The authors have declared no competing interest.

### Author Declarations

Ethical approval for the HELIUS study was obtained from the Academic Medical Center Ethical Review Board. All participants provided written informed consent.

