## Supplement for "SARS-CoV-2 antibody prevalence and determinants of six ethnic groups living in Amsterdam, the Netherlands: a population-based cross-sectional study, June-October 2020"

**Content** **Page**

Information on seroprevalence estimation corrected for sampling, post-stratification and adjusting for differences in age, sex and calendar time between ethnic groups. 2

Table S1. Characteristics of three inclusion groups (invited and included in SARS-CoV-2 serological study invited not included not invited) within the HELIUS population (N=16889), Amsterdam, the Netherlands, 24 June - 9 October 2020 3

Figure S1 Inclusion numbers and test results per month by ethnicity, Amsterdam, the Netherlands, 24 June - 9 October 2020 5

Figure S2 Distribution of qualitative signal-to-cutoff (S/CO) ratios for positive test results (N=226) by ethnicity, Amsterdam, the Netherlands, 24 June - 9 October 2020 6

*Univariable analysis of potential determinants of SARS-CoV-2 seropositivity:*

Table S2: in Dutch participants 7

Table S3: in South-Asian Surinamese participants 10

Table S4: in African Surinamese participants 12

Table S5: in Ghanaian participants 16

Table S6: in Turkish participants 19

Table S7: in Moroccan participants 22

**Information on seroprevalence estimation corrected for sampling, post-stratification and adjusting for differences in age, sex and calendar time between ethnic groups.**

For sampling, the probability of being invited for the COVID-19 substudy (as the proportion of participants invited among those in active follow-up in the parent study) was calculated, as was the conditional probability of participating in the COVID-19 substudy (given the participant's ethnicity, age, educational level, working status and health literacy). The product of the two probabilities was taken and the inverse of this result, standardized to one, was used as a sampling weight. For post-stratification, a weight was assigned corresponding to the proportion representing the Amsterdam population of each stratum of age (20-44, 45-54, 55-59, 60-79 years), sex (male, female) and ethnicity (Surinamese, Ghanaian, Moroccan, Turkish, Dutch). Sampling and post-stratification weights were placed in a multivariable logistic regression model with covariates ethnicity, age, sex, and calendar time. Given the weighting scheme of this study, variance was calculated with the designed-based Taylor series linearization method using the 'svy' commands in STATA. Differences between ethnic groups were tested in the model using the Wald $\chi^2$ test.

**Table S1. Characteristics of three inclusion groups (invited and included in COVID-19 study invited not included not invited) within the HELIUS population (N=16889), Amsterdam, the Netherlands, 24 June - 9 October 2020**

To identify potential selection bias among HELIUS participants who were still in active follow-up, demographic, socio-economic factors and access to health care indicators were compared between those who were invited versus not invited for the COVID-19 substudy. To assess the reasons for nonresponse among invited HELIUS participants, these variables were also compared between those who participated versus not participated in the COVID-19 substudy. Pearson's χ2 or Fisher exact test were used for categorical data and Kruskal-Wallis rank test for continuous variables.

| Characteristic | All HELIUS participants in follow-up[a] (N= 16889) | Invited included (n=2497) | Invited not included (n=8583) | Not invited (n=5809) | *Invited and included vs. invited not included* | *Invited (included and not included) vs. not invited* |
|---|---|---|---|---|---|---|
| | n (%) | n (%) | n (%) | n (%) | *P*-value | *P*-value |
| **Ethnicity** | | | | | <0.001 | <0.001 |
| Dutch | 3029 (17.9%) | 503 (20.1%) | 506 (5.9%) | 2020 (34.8%) | | |
| South-Asian Surinamese | 2328 (13.8%) | 453 (18.1%) | 1088 (12.7%) | 787 (13.5%) | | |
| African Surinamese | 2895 (17.1%) | 407 (16.3%) | 1103 (12.9%) | 1385 (23.8%) | | |
| Ghanaian | 2166 (12.8%) | 331 (13.3%) | 1832 (21.3%) | 3 (0.1%) | | |
| Turkish | 3071 (18.2%) | 409 (16.4%) | 2162 (25.2%) | 500 (8.6%) | | |
| Moroccan | 3400 (20.1%) | 394 (15.8%) | 1892 (22.0%) | 1114 (19.2%) | | |
| **Sex** | | | | | 0.095 | 0.94 |
| Male | 7077 (41.9%) | 1083 (43.4%) | 3562 (41.5%) | 2432 (41.9%) | | |
| Female | 9812 (58.1%) | 1414 (56.6%) | 5021 (58.5%) | 3377 (58.1%) | | |
| **Age in years on 1 January 2020** | | | | | <0.001 | <0.001 |
| Median [IQR] | 52 [41-61] | 54 [44-61] | 51 [39-59] | 54 [42-63] | | |
| **Migration generation** | | | | | <0.001 | <0.001 |
| N.A. (Dutch group) | 3029 (17.9%) | 503 (20.1%) | 506 (5.9%) | 2020 (34.8%) | | |
| 1st | 10978 (65.0%) | 1656 (66.3%) | 6339 (73.9%) | 2983 (51.4%) | | |
| 2nd | 2882 (17.1%) | 338 (13.5%) | 1738 (20.2%) | 806 (13.9%) | | |
| **City district[b]** | | | | | <0.001 | <0.001 |
| Centre | 783 (4.6%) | 142 (5.7%) | 222 (2.6%) | 419 (7.2%) | | |
| East | 2550 (15.1%) | 421 (16.9%) | 1302 (15.2%) | 827 (14.2%) | | |
| West | 2361 (14.0%) | 297 (11.9%) | 1205 (14.0%) | 859 (14.8%) | | |
| South | 1382 (8.2%) | 248 (9.9%) | 524 (6.1%) | 610 (10.5%) | | |
| New-West | 4893 (29.0%) | 603 (24.1%) | 2571 (30.0%) | 1719 (29.6%) | | |
| Southeast | 4803 (28.4%) | 765 (30.6%) | 2722 (31.7%) | 1316 (22.7%) | | |
| Other | 16 (0.1%) | 3 (0.1%) | 7 (0.1%) | 6 (0.1%) | | |
| Missing | 101 (0.6%) | 18 (0.7%) | 30 (0.3%) | 53 (0.9%) | | |
| **Educational level[b]** | | | | | <0.001 | <0.001 |
| No school/elementary school | 3286 (19.5%) | 327 (13.1%) | 2175 (25.3%) | 784 (13.5%) | | |
| Lower vocational/ lower secondary school | 4324 (25.6%) | 612 (24.5%) | 2358 (27.5%) | 1354 (23.3%) | | |
| Intermediary vocational/ intermediary secondary school | 4715 (27.9%) | 700 (28.0%) | 2393 (27.9%) | 1622 (27.9%) | | |

| | | | | | | |
|---|---|---|---|---|---|---|
| Higher vocational/university | 3993 (23.6%) | 792 (31.7%) | 1243 (14.5%) | 1958 (33.7%) | | |
| Missing | 571 (3.4%) | 66 (2.6%) | 414 (4.8%) | 91 (1.6%) | | |
| **Labor participation**[b] | | | | | <0.001 | <0.001 |
| Employed | 9585 (56.8%) | 1659 (66.4%) | 4274 (49.8%) | 3652 (62.9%) | | |
| Not in workforce | 2992 (17.7%) | 309 (12.4%) | 1645 (19.2%) | 1038 (17.9%) | | |
| Unemployed/on benefits | 2372 (14.0%) | 300 (12.0%) | 1416 (16.5%) | 656 (11.3%) | | |
| Disabled | 1309 (7.8%) | 151 (6.0%) | 792 (9.2%) | 366 (6.3%) | | |
| Missing | 631 (3.7%) | 130 (3.1%) | 774 (8.7%) | 154 (2.7%) | | |
| **Occupational level**[b] | | | | | <0.001 | <0.001 |
| Elementary occupations | 2454 (14.5%) | 323 (12.9%) | 1739 (20.3%) | 392 (6.7%) | | |
| Lower occupations | 4177 (24.7%) | 537 (21.5%) | 2280 (26.6%) | 1360 (23.4%) | | |
| Intermediary occupations | 3549 (21.0%) | 599 (24.0%) | 1515 (17.7%) | 1435 (24.7%) | | |
| Higher occupations | 2565 (15.2%) | 500 (20.0%) | 783 (9.1%) | 1282 (22.1%) | | |
| Scientific occupations | 928 (5.5%) | 202 (8.1%) | 223 (2.6%) | 503 (8.7%) | | |
| Missing | 3216 (19.0%) | 336 (13.5%) | 2043 (23.8%) | 837 (14.4%) | | |
| **Difficulty with Dutch language**[b] | | | | | <0.001 | <0.001 |
| N.A. (Dutch group) | 3029 (17.9%) | 503 (20.1%) | 506 (5.9%) | 2020 (34.8%) | | |
| No | 7467 (44.2%) | 1148 (46.0%) | 3751 (43.7%) | 2568 (44.2%) | | |
| Yes | 5891 (34.9%) | 782 (31.3%) | 3950 (46.0%) | 1159 (20.0%) | | |
| Missing | 502 (3.0%) | 64 (2.6%) | 376 (4.4%) | 62 (1.1%) | | |
| **Difficulty with Dutch language**[b] **(excluding Dutch group)** | | | | | <0.001 | <0.001 |
| No | 7467 (53.9%) | 1148 (57.6%) | 3751 (46.4%) | 2568 (67.8%) | | |
| Yes | 5891 (42.5%) | 782 (39.2%) | 3950 (48.9%) | 1159 (30.6%) | | |
| Missing | 502 (3.6%) | 64 (3.2%) | 376 (4.7%) | 62 (1.6%) | | |
| **Health literacy (SBSQ)**[b] | | | | | <0.001 | <0.001 |
| Adequate | 5329 (31.6%) | 971 (38.9%) | 2058 (24.0%) | 2300 (39.6%) | | |
| Low | 1927 (11.4%) | 265 (10.6%) | 1110 (12.9%) | 552 (9.5%) | | |
| Missing | 9633 (57.0%) | 1261 (50.5%) | 5415 (63.1%) | 2957 (50.9%) | | |

**Abbreviations:** HELIUS Healthy Life in an Urban Setting; IQR interquartile range; N.A. not applicable; SBSQ Set of Brief Screening Question

[a] Excluding participants not belonging to one of the six ethnic groups included in the COVID-19 study [b] Measured at baseline (2011-2015)

**Figure S1 Inclusion numbers and test results per month by ethnicity, Amsterdam, the Netherlands, 24 June - 9 October 2020**

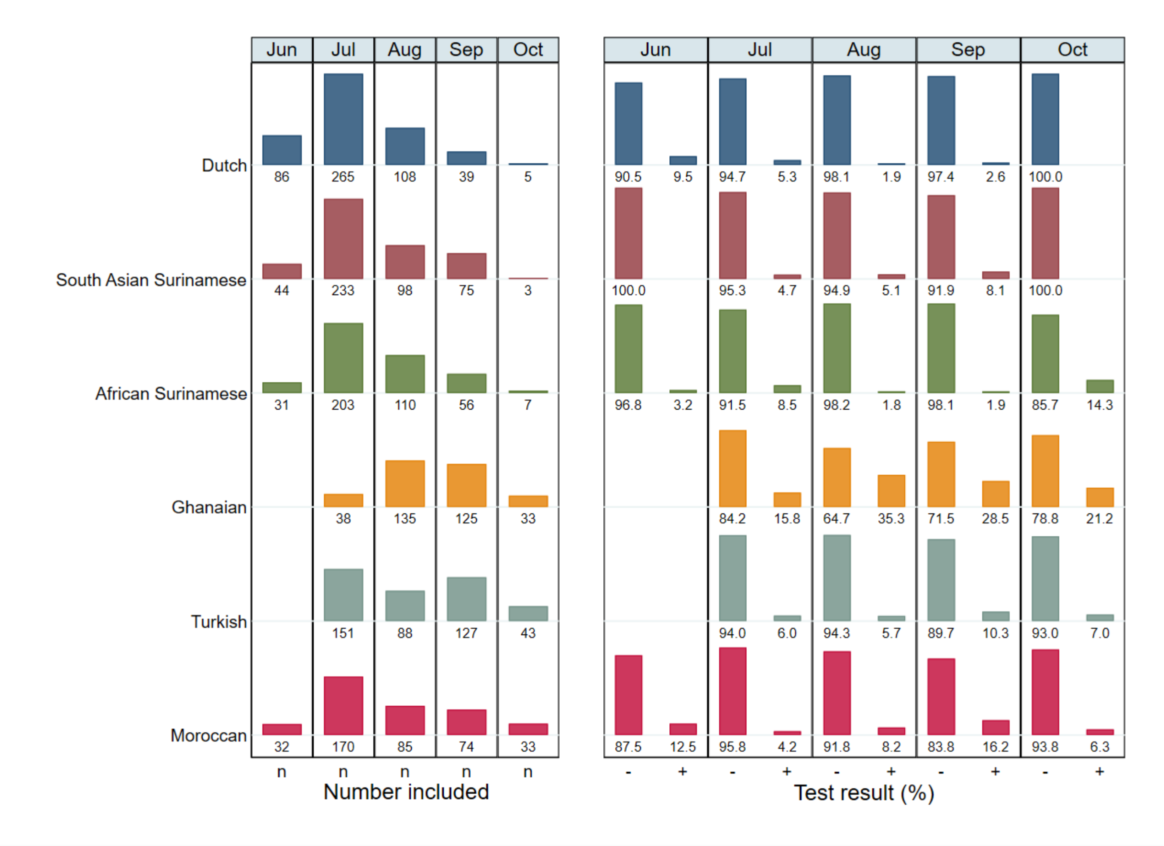


The left side of the graph shows the number of individuals included in the substudy per month by ethnic group. The right side of the graph shows the distribution of test results per inclusion month by ethnic group, excluding people without a test result (n=14) or equivocal test result (n=8).

We tested whether the seroprevalence changed over months in survey-weighted logistic regression models per ethnic group. Odds of a positive test did not change in the Dutch (*P*=0.91), Ghanaian (*P*=0.33), Turkish (*P*=0.67) and Moroccan groups (*P*=0.33), but increased in the South-Asian Surinamese group (OR=1.87 per month increase, 95%CI=1.12-3.12, *P*=0.016) and decreased in the African Surinamese group (OR=0.56 per month increase, 95%CI=0.34-0.94, *P*=0.028).

**Figure S2 Distribution of signal-to-cutoff (S/CO) ratios for positive test results (N=226) by ethnicity, Amsterdam, the Netherlands, 24 June - 9 October 2020**

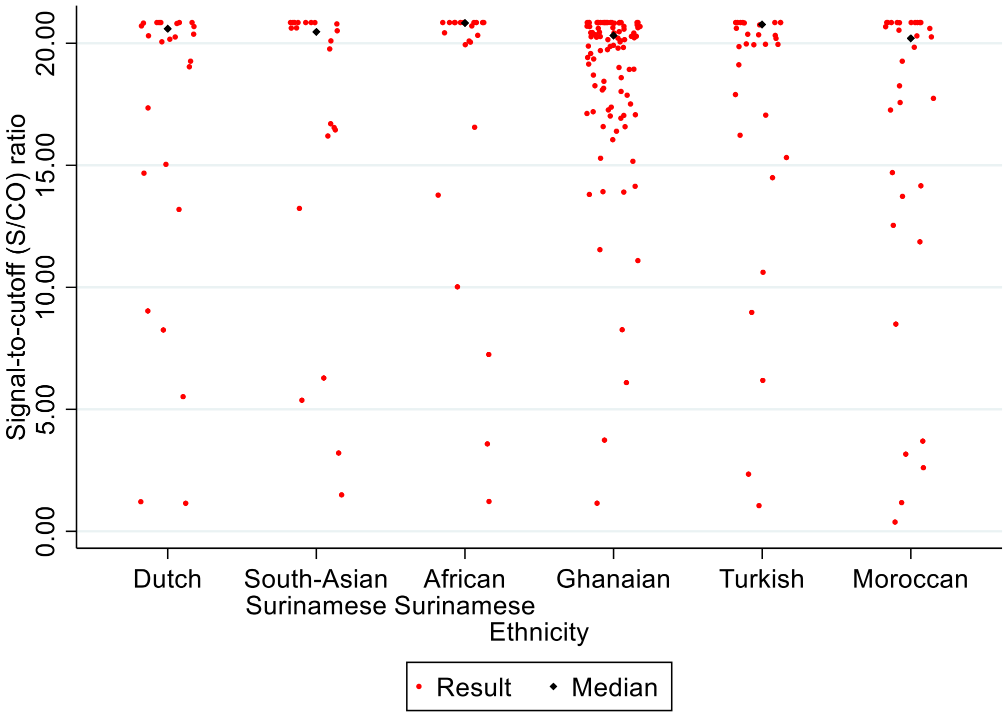


Kruskall Wallis test for difference between ethnic groups: *P*=0.50

**Table S2. Univariable analysis of potential determinants of SARS-CoV-2 seropositivity in Dutch participants, Amsterdam, the Netherlands, 24 June - 9 October 2020**

| Characteristic | Number per category | Number with antibodies (%) | OR (95% CI)[a] | *P*-value |
|---|---|---|---|---|
| **Sex** | | | | 0.26 |
| Male | 234 | 10 (4.3%) | 1 | |
| Female | 264 | 15 (5.7%) | 1.79 (0.65-4.88) | |
| **Per year increase in age in years on 1 January 2020**[c] | | | 0.97 (0.93-1.00) | 0.087 |
| **Migration generation** | | | | |
| 1st | N.A. | N.A. | | |
| 2nd | N.A. | N.A. | | |
| **Month of study visit**[b] | | | | 0.55 |
| June | 84 | 8 (9.5%) | 1 | |
| July | 262 | 14 (5.3%) | 0.78 (0.26-2.33) | |
| August | 108 | 2 (1.9%) | 0.32 (0.06-1.63) | |
| September | 39 | 1 (2.6%) | 0.46 (0.05-4.07) | |
| October | 5 | 0 (0%) | Omitted | |
| **City district**[c] | | | | 0.88 |
| Centre | 86 | 7 (8.1%) | 1 | |
| East | 99 | 2 (2.0%) | 1.11 (0.21-5.92) | |
| West | 89 | 5 (5.6%) | 1.11 (0.27-4.63) | |
| South | 112 | 8 (7.1%) | 1.78 (0.58-5.49) | |
| New-West | 44 | 3 (6.8%) | 1.64 (0.30-8.85) | |
| Southeast | 63 | 0 (0%) | Omitted | |
| Other | 1 | 0 (0%) | Omitted | |
| **Has obesity (BMI≥30.0)**[c] | | | | 0.66 |
| No | 442 | 22 (5.0%) | 1 | |
| Yes | 51 | 3 (5.9%) | 0.75 (0.21-2.72) | |
| **Educational level**[c] | | | | 0.32 |
| No school/elementary school | 10 | 0 (0%) | Omitted | |
| Lower vocational/ lower secondary school | 53 | 0 (0%) | Omitted | |
| Intermediary vocational/ intermediary secondary school | 99 | 1 (1.0%) | 1 | |
| Higher vocational/university | 335 | 24 (7.2%) | 2.78 (0.37-20.97) | |
| Missing | 1 | 0 (0%) | Omitted | |
| **Labor participation**[c] | | | | 0.19 |
| Employed | 377 | 19 (5.0%) | 1 | |
| Not in workforce | 88 | 6 (6.8%) | 2.12 (0.69-6.55) | |
| Unemployed/on benefits | 21 | 0 (0%) | Omitted | |
| Disabled | 11 | 0 (0%) | Omitted | |
| Unknown/missing | 1 | 0 (0%) | Omitted | |
| **Elementary occupation**[c] | | | | 0.20 |
| No | 467 | 23 (4.9%) | 1 | |
| Yes | 4 | 0 (0%) | Omitted | |
| Missing | 27 | 2 (7.4) | 2.71 (0.58-12.64) | |
| **Health literacy (SBSQ)**[c] | | | | |
| Adequate | 495 | 25 (5.1%) | Omitted | |
| Low | 3 | 0 (0%) | Omitted | |
| **Job setting**[b,e] | | | | |
| No job / caretaker only | 115 | 2 (1.7%) | 1 | |

| | | | | |
|---|---|---|---|---|
| Job with no contact within 1.5 meter | 96 | 3 (3.1%) | 3.40 (0.44-25.99) | |
| Other job with contact within 1.5 meter | 142 | 10 (7.0%) | 6.22 (1.25-30.86) | |
| Child care/schools/higher education | 62 | 4 (6.5%) | 8.23 (1.26-53.64) | |
| Bar/restaurant | 12 | 1 (8.3%) | 2.51 (0.20-32.41) | |
| Hospital/long-term care facility/Care worker elsewhere | 71 | 5 (7.0%) | 8.51 (1.37-52.99) | |
| **Caretaker** [b] | | | | 0.46 |
| No | 424 | 20 (4.7%) | 1 | |
| Yes | 74 | 5 (6.8) | 1.68 (0.43-6.6) | |
| **Number of people in household** [c] | | | | 0.067 |
| 1 (Lives alone) | 125 | 6 (4.8%) | 1 | |
| 2 | 222 | 12 (5.4%) | 0.61 (0.18-2.06) | |
| 3 | 63 | 1 (1.6%) | 0.07 (0.01-0.63) | |
| 4 | 76 | 4 (5.3%) | 0.56 (0.14-2.31) | |
| ≥5 | 12 | 2 (16.7%) | 3.45 (0.44-27.16) | |
| **Lives with other people** [b] | 375 | 20 (5.3%) | 0.76 (0.22-2.61) | 0.66 |
| **Partner** | 334 | 19 (5.7%) | 0.79 (0.26-2.41) | 0.68 |
| **Children up to 3 years old** | 38 | 1 (2.6%) | 0.36 (0.05-2.85) | 0.33 |
| **Children 4 through 12 years old** | 55 | 2 (3.6%) | 0.52 (0.11-2.43) | 0.41 |
| **Children 13 through 17 years old** | 31 | 0 (0%) | Omitted | |
| **Children 18+ years old** | 50 | 1 (2.0%) | 0.23 (0.03-1.77) | 0.16 |
| **Parents or parents-in-law** | 4 | 0 (0%) | Omitted | |
| **Other adults** | 18 | 1 (5.6%) | 2.17 (0.26-18.01) | 0.47 |
| **Household member/steady partner with suspected infection** [b] | | | | 0.001 |
| N.A./No | 440 | 12 (2.7%) | 1 | |
| Yes | 53 | 13 (24.5%) | 6.26 (2.16-18.13) | |
| **Number of times left home in the past week** [b,d] | | | | 0.19 |
| 0-7 | 59 | 1 (1.7%) | 1 | |
| 8-11 | 81 | 4 (4.9%) | 4.22 (0.42-42.42) | |
| 12-16 | 140 | 6 (4.3%) | 4.51 (0.44-46.04) | |
| 17+ | 218 | 14 (6.4%) | 8.42 (1.07-66.63) | |
| **In the past week, left home to** [b]**:** | | | | |
| **Work** | 237 | 16 (6.8%) | 1.72 (0.60-4.93) | 0.32 |
| **Do groceries** | 473 | 23 (4.9%) | 1.64 (0.35-7.72) | 0.53 |
| **Visit family or friends** | 335 | 20 (6.0%) | 1.68 (0.48-5.93) | 0.42 |
| **Walk the dog or go outside with kids** | 125 | 8 (6.4%) | 1.34 (0.46-3.85) | 0.59 |
| **Walk or exercise outside** | 396 | 19 (4.8%) | 1.01 (0.30-3.39) | 0.99 |
| **Take care of someone** | 75 | 6 (8.0%) | 1.12 (0.39-3.20) | 0.83 |
| **Pick up prescription medicines or visit** | 99 | 10 (10.1%) | 3.97 (1.38-11.43) | 0.01 |
| **Attend religious service** | 5 | 0 (0%) | Omitted | |
| **Visit cultural place** | 80 | 5 (6.3%) | 1.24 (0.38-4.02) | 0.72 |
| **Visit bar or restaurant** | 279 | 15 (5.4%) | 1.61 (0.59-4.35) | 0.35 |
| **Indoor sports** | 63 | 4 (6.3%) | 1.56 (0.40-6.11) | 0.52 |
| **Visit recreational park** | 210 | 11 (5.2%) | 1.01 (0.36-2.83) | 0.98 |
| **Frequency of using public transportation in the past week** [b] | | | | 0.79 |
| 0 days | 321 | 17 (5.3%) | 1 | |
| 1-2 days | 130 | 6 (4.6%) | 0.64 (0.18-2.32) | |
| 3-4 days | 36 | 2 (5.6%) | 0.84 (0.14-4.87) | |
| 5-7 days | 10 | 0 (0%) | Omitted | |
| **Number of unique visitors at home in the past week** [b] | | | | 0.50 |

| | | | | |
|---|---|---|---|---|
| 0 | 213 | 9 (4.2%) | 1 | |
| 1 | 88 | 4 (4.5%) | 1.21 (0.24-6.24) | |
| 2-4 | 145 | 6 (4.1%) | 0.72 (0.21-2.44) | |
| 5+ | 49 | 6 (12.2%) | 2.20 (0.56-8.71) | |
| **Travelled abroad in 2020** [b] | | | | 0.021 |
| No | 259 | 8 (3.1%) | 1 | |
| Yes | 239 | 17 (7.1%) | 3.43 (1.21-9.75) | |

**Abbreviations:** CI, confidence interval; HELIUS, Healthy Life in an Urban Setting; N.A., not applicable; OR, odds ratio
[a] Those with an equivocal test result were excluded from this analysis [b] Measured at COVID-1 visit (2020) [c] Measured at baseline (2011-2015) [d] Quartiles [e] Presumed higher exposure categories had priority, i.e. if someone was working in a school and as a careworker, they were categorized as a health worker. Caretakers were not included as a category because many had other jobs.

In multivariable analysis, the distribution of educational level and labor participation were skewed to mostly one group and hence were not included. The following variables were removed as they were no longer significant in the multivariable model: dichotomized household size ($P$=0.80), age (0.53), occupational level (0.36), number of times left home (0.40), living with child 18+ years old (0.19), job setting (0.12).

**Table S3. Univariable analysis of potential determinants of SARS-CoV-2 seropositivity in South-Asian Surinamese participants, Amsterdam, the Netherlands, 24 June - 9 October 2020**

| Characteristic | Number per category | Number with antibodies (%) | OR (95% CI)[a] | *P*-value |
|---|---|---|---|---|
| **Sex** | | | | 0.84 |
| Male | 178 | 7 (3.9%) | 1 | |
| Female | 273 | 15 (5.5%) | 1.13 (0.34-3.77) | |
| **Per year increase in age in years on 1 January 2020**[c] | | | 0.98 (0.95-1.02) | 0.44 |
| **Migration generation** | | | | 0.36 |
| 1st | 368 | 17 (4.6%) | 1 | |
| 2nd | 83 | 5 (6.0%) | 1.68 (0.56-5.05) | |
| **Month of study visit**[b] | | | | 0.16 |
| June | 44 | 0 (0%) | Omitted | |
| July | 232 | 11 (4.7%) | 1 | |
| August | 98 | 5 (5.1%) | 1.79 (0.46-7.04) | |
| September | 74 | 6 (8.1%) | 3.22 (0.95-10.91) | |
| October | 3 | 0 (0%) | Omitted | |
| **City district**[c] | | | | 0.48 |
| Centre | 17 | 0 (0%) | Omitted | |
| East | 53 | 2 (3.8%) | 1 | |
| West | 6 | 0 (0%) | Omitted | |
| South | 32 | 1 (3.1%) | 1.05 (0.09-12.53) | |
| New-West | 111 | 5 (4.5%) | 1.43 (0.25-8.24) | |
| Southeast | 228 | 14 (6.1%) | 2.81 (0.56-14.10) | |
| Other | 0 | 0 (0%) | Omitted | |
| **Has obesity (BMI≥30.0)**[c] | | | | 0.55 |
| No | 369 | 20 (5.4%) | 1 | |
| Yes | 76 | 2 (2.6%) | 0.58 (0.10-3.42) | |
| **Educational level**[c] | | | | 0.62 |
| No school/elementary school | 55 | 2 (3.6%) | 1 | |
| Lower vocational/ lower secondary school | 156 | 11 (7.1%) | 2.64 (0.53-13.21) | |
| Intermediary vocational/ intermediary secondary school | 136 | 5 (3.7%) | 1.41 (0.22-9.14) | |
| Higher vocational/university | 103 | 4 (3.9%) | 2.06 (0.28-14.89) | |
| Missing | 1 | 0 (0%) | Omitted | |
| **Labor participation**[c] | | | | 0.53 |
| Employed | 306 | 16 (5.2%) | 1 | |
| Not in workforce | 47 | 1 (2.1%) | 0.84 (0.1-6.77) | |
| Unemployed/on benefits | 53 | 3 (5.7%) | 2.66 (0.56-12.59) | |
| Disabled | 39 | 1 (2.6%) | 0.55 (0.07-4.38) | |
| Unknown/missing | 6 | 1 (16.7%) | 3.43 (0.37-31.95) | |
| **Elementary occupation**[c] | | | | 0.12 |
| No | 367 | 18 (4.9%) | 1 | |
| Yes | 36 | 3 (8.3%) | 1.19 (0.30-4.69) | |
| Missing | 48 | 1 (2.1%) | 0.12 (0.02-0.98) | |
| **Difficulty with Dutch language**[c] | | | | 0.48 |
| No | 347 | 13 (3.7%) | 1 | |
| Yes | 103 | 9 (8.7%) | 1.45 (0.52-4.04) | |
| **Health literacy (SBSQ)**[c] | | | | 0.94 |
| Adequate | 435 | 21 (4.8%) | 1 | |

| | | | | |
|---|---|---|---|---|
| Low | 16 | 1 (6.3%) | 0.93 (0.11-7.8) | |
| **Job setting[b,e]** | | | | 0.06 |
| No job / caretaker only | 144 | 5 (3.5%) | 1 | |
| Job with no contact within 1.5 meter | 65 | 1 (1.5%) | 0.27 (0.03-2.42) | |
| Other job with contact within 1.5 meter | 153 | 12 (7.8%) | 3.35 (0.99-11.32) | |
| Child care/schools/higher education | 27 | 1 (3.7%) | 1.19 (0.12-11.38) | |
| Bar/restaurant | 10 | 1 (10.0%) | 1.28 (0.12-13.30) | |
| Hospital/long-term care facility/Care worker elsewhere | 50 | 2 (4.0%) | 0.46 (0.08-2.61) | |
| **Caretaker [b]** | | | 0.27 (0.03-2.42) | |
| No | 380 | 20 (5.3%) | 3.35 (0.99-11.32) | |
| Yes | 69 | 2 (2.9%) | 1.19 (0.12-11.38) | |
| **Number of people in household [c]** | | | | 0.02 |
| 1 (Lives alone) | 98 | 1 (1.0%) | 1 | |
| 2 | 126 | 7 (5.6%) | 4.55 (0.53-39.15) | |
| 3 | 102 | 10 (9.8%) | 16.85 (1.99-142.58) | |
| 4 | 77 | 3 (3.9%) | 2.96 (0.30-29.11) | |
| ≥5 | 44 | 1 (2.3%) | 1.69 (0.10-28.05) | |
| **Lives with other people [b]** | 333 | 18 (5.4%) | 1.91 (0.57-6.46) | 0.29 |
| **Partner** | 224 | 13 (5.8%) | 1.11 (0.35-3.47) | 0.86 |
| **Children up to 3 years old** | 19 | 1 (5.3%) | 0.91 (0.11-7.49) | 0.93 |
| **Children 4 through 12 years old** | 51 | 0 (0%) | Omitted | |
| **Children 13 through 17 years old** | 40 | 2 (5.0%) | 0.68 (0.14-3.24) | 0.63 |
| **Children 18+ years old** | 146 | 7 (4.8%) | 0.85 (0.30-2.42) | 0.76 |
| **Parents or parents-in-law** | 32 | 3 (9.4%) | 2.03 (0.50-8.19) | 0.32 |
| **Other adults** | 32 | 0 (0%) | Omitted | |
| **Household member/steady partner with suspected infection [b]** | | | | 0.002 |
| N.A./No | 408 | 13 (3.2%) | 1 | |
| Yes | 38 | 9 (23.7%) | 7.05 (2.07-24.04) | |
| **Number of times left home in the past week [b,d]** | | | | 0.02 |
| 0-7 | 144 | 9 (6.3%) | 1 | |
| 8-11 | 134 | 9 (6.7%) | 2.12 (0.64-6.98) | |
| 12-16 | 102 | 2 (2.0%) | 0.18 (0.04-0.94) | |
| 17+ | 69 | 2 (2.9%) | 0.49 (0.09-2.56) | |
| **In the past week, left home to [b]:** | | | | |
| **Work** | 195 | 9 (4.6%) | 0.62 (0.19-2.09) | 0.44 |
| **Do groceries** | 422 | 20 (4.7%) | 1.28 (0.25-6.56) | 0.77 |
| **Visit family or friends** | 247 | 12 (4.9%) | 1.01 (0.34-3.03) | 0.98 |
| **Walk the dog or go outside with kids** | 34 | 1 (2.9%) | 0.55 (0.07-4.46) | 0.58 |
| **Walk or exercise outside** | 266 | 12 (4.9%) | 1.86 (0.68-5.03) | 0.22 |
| **Take care of someone** | 63 | 1 (2.9) | 0.23 (0.03-1.79) | 0.16 |
| **Pick up prescription medicines or visit** | 120 | 12 (4.5%) | 1.77 (0.57-5.54) | 0.33 |
| **Attend religious service** | 20 | 1 (1.6%) | 0.73 (0.09-6.09) | 0.77 |
| **Visit cultural place** | 17 | 7 (5.8%) | 0.89 (0.1-7.57) | 0.91 |
| **Visit bar or restaurant** | 87 | 1 (5.0%) | 0.48 (0.06-3.79) | 0.49 |
| **Indoor sports** | 77 | 4 (5.2%) | 1.26 (0.29-5.47) | 0.75 |
| **Visit recreational park** | 66 | 3 (4.5%) | 1.45 (0.31-6.76) | 0.64 |
| **Frequency of using public transportation in the past week [b]** | | | | 0.95 |
| 0 days | 316 | 16 (5.1%) | 1 | |
| 1-2 days | 76 | 3 (3.9%) | 0.81 (0.20-3.26) | |
| 3-4 days | 29 | 0 (0%) | Omitted | |

| | | | | |
|---|---|---|---|---|
| 5-7 days | 27 | 3 (11.1%) | 1.06 (0.25-4.42) | |
| **Number of unique visitors at home in the past week** [b] | | | | 0.27 |
| 0 | 217 | 12 (5.5%) | 1 | |
| 1 | 80 | 2 (2.5%) | 0.37 (0.07-1.92) | |
| 2-4 | 119 | 6 (5.0%) | 1.07 (0.31-3.68) | |
| 5+ | 30 | 2 (6.7%) | 3.68 (0.53-25.58) | |
| **Travelled abroad in 2020** [b] | | | | 0.010 |
| No | 332 | 15 (4.5%) | 1 | |
| Yes | 117 | 7 (6.0%) | 4.06 (1.40-11.76) | |

**Abbreviations:** CI, confidence interval; HELIUS, Healthy Life in an Urban Setting; OR, odds ratio

[a] Those with an equivocal test result were excluded from this analysis [b] Measured at COVID-1 visit (2020) [c] Measured at baseline (2011-2015) [d] Quartiles [e] Presumed higher exposure categories had priority, i.e. if someone was working in a school and as a careworker, they were categorized as a health worker. Caretakers were not included as a category because many had other jobs.

In multivariable analysis, the distribution of occupational level and number of times left home were skewed to mostly one group and hence were not included. The following variables were removed as they were no longer significant in the multivariable model: job setting (*P*=0.84), leaving home to care for someone (0.32), else dichotomized household size (0.18).

**Table S4. Univariable analysis of potential determinants of SARS-CoV-2 seropositivity in African Surinamese participants, Amsterdam, the Netherlands, 24 June - 9 October 2020**

| Characteristic | Number per category | Number with antibodies (%) | OR (95% CI)[a] | *P*-value |
|---|---|---|---|---|
| **Sex** | | | | 0.70 |
| Male | 163 | 7 (4.3%) | 1 | |
| Female | 237 | 15 (6.3%) | 0.76 (0.20-2.98) | |
| **Per year increase in age in years on 1 January 2020[c]** | | | 0.94 (0.88-1.00) | 0.063 |
| **Migration generation** | | | | 0.030 |
| 1st | | | 1 | |
| 2nd | | | 3.97 (1.11-14.28) | |
| **Month of study visit[b]** | | | | 0.020 |
| June | 31 | 1 (3.2%) | 1 | |
| July | 199 | 17 (8.5%) | 4.80 (0.57-40.24) | |
| August | 109 | 2 (1.8%) | 0.35 (0.03-4.03) | |
| September | 54 | 1 (1.9%) | 0.83 (0.05-13.72) | |
| October | 7 | 1 (14.3%) | 3.42 (0.18-64.59) | |
| **City district [c]** | | | | 0.50 |
| Centre | 15 | 1 (6.7%) | 1 | |
| East | 81 | 5 (6.2%) | 0.93 (0.10-8.7) | |
| West | 34 | 1 (2.9%) | 0.25 (0.02-4.36) | |
| South | 28 | 1 (3.6%) | 2.04 (0.12-35.25) | |
| New-West | 49 | 3 (6.1%) | 0.92 (0.09-9.58) | |
| Southeast | 190 | 11 (5.8%) | 2.04 (0.22-19.04) | |
| Other | 1 | 0 (0%) | Omitted | |
| **Has obesity (BMI≥30.0) [c]** | | | | 0.88 |
| No | 312 | 15 (4.8%) | 1 | |
| Yes | 87 | 7 (8.0%) | 0.92 (0.3-2.81) | |
| **Educational level [c]** | | | | 0.78 |
| No school/elementary school | 14 | 1 (7.1%) | 1 | |
| Lower vocational/ lower secondary school | 118 | 7 (5.9%) | 2.66 (0.24-28.99) | |
| Intermediary vocational/ intermediary secondary school | 142 | 9 (6.3%) | 1.54 (0.16-14.82) | |
| Higher vocational/university | 124 | 5 (4.0%) | 1.22 (0.12-12.53) | |
| Missing | 2 | 0 (0%) | Omitted | |
| **Labor participation [c]** | | | | 0.016 |
| Employed | 289 | 14 (4.8%) | 1 | |
| Not in workforce | 37 | 4 (10.8%) | 8.09 (1.85-35.42) | |
| Unemployed/on benefits | 46 | 2 (4.3%) | 0.41 (0.09-2.02) | |
| Disabled | 24 | 2 (8.3%) | 1.26 (0.25-6.47) | |
| Unknown/missing | 3 | 0 (0%) | Omitted | |
| **Elementary occupation [c]** | | | | 0.081 |
| No | 350 | 18 (5.1%) | 1 | |
| Yes | 22 | 2 (9.1%) | 1.83 (0.38-8.82) | |
| Missing | 28 | 2 (7.1%) | 6.64 (1.25-35.31) | |
| **Difficulty with Dutch language [c]** | | | | 0.21 |
| No | 353 | 20 (5.7%) | 1 | |
| Yes | 45 | 2 (4.4%) | 0.36 (0.07-1.78) | |
| **Health literacy (SBSQ) [c]** | | | | 0.98 |
| Adequate | 393 | 21 (5.3%) | 1 | |
| Low | 7 | 1 (14.3%) | 1.03 (0.10-10.43) | |

| | | | | |
|---|---|---|---|---|
| **Job setting[b,e]** | | | | 0.046 |
| No job / caretaker only | 117 | 5 (4.3%) | 1 | |
| Job with no contact within 1.5 meter | 39 | 1 (2.6%) | 0.21 (0.02-1.93) | |
| Other job with contact within 1.5 meter | 130 | 7 (5.4%) | 2.20 (0.48-10.06) | |
| Child care/schools/higher education | 42 | 1 (2.4%) | 0.31 (0.03-2.78) | |
| Bar/restaurant | 11 | 0 (0%) | Omitted | |
| Hospital/long-term care facility/Care worker elsewhere | 61 | 8 (13.1%) | 3.09 (0.81-11.7) | |
| **Caretaker [b]** | | | | 0.81 |
| No | 336 | 18 (5.4%) | 1 | |
| Yes | 64 | 4 (6.3%) | 0.85 (0.23-3.14) | |
| **Number of people in household [c]** | | | | |
| 1 (Lives alone) | 143 | 2 (1.4%) | 1 | 0.039 |
| 2 | 88 | 7 (8.0%) | 12.95 (2.21-76.01) | |
| 3 | 75 | 5 (6.7%) | 17.30 (2.45-122.24) | |
| 4 | 63 | 5 (7.9%) | 6.26 (1.11-35.42) | |
| ≥5 | 26 | 3 (11.5%) | 8.09 (1.19-55.04) | |
| **Lives with other people [b]** | 255 | 18 (7.1%) | 2.19 (0.43-11.24) | 0.35 |
| **Partner** | 163 | 11 (6.7%) | 0.78 (0.23-2.66) | 0.69 |
| **Children up to 3 years old** | 19 | 2 (10.5%) | 2.09 (0.38-11.54) | 0.40 |
| **Children 4 through 12 years old** | 44 | 1 (2.3%) | 0.12 (0.01-0.95) | 0.039 |
| **Children 13 through 17 years old** | 41 | 2 (4.9%) | 0.32 (0.07-1.58) | 0.16 |
| **Children 18+ years old** | 110 | 11 (10.0%) | 1.22 (0.41-3.64) | 0.72 |
| **Parents or parents-in-law** | 11 | 2 (18.2%) | 1.63 (0.3-9.04) | 0.57 |
| **Other adults** | 27 | 4 (14.8%) | 9.34 (1.7-51.41) | 0.010 |
| **Household member/steady partner with suspected infection [b]** | | | | <0.001 |
| N.A./No | 367 | 11 (3.0%) | 1 | |
| Yes | 33 | 11 (33.3%) | 20.08 (4.98-80.9) | |
| **Number of times left home in the past week [b,d]** | | | | 0.38 |
| 0-7 | 143 | 10 (7.0%) | 1.52 (0.32-7.21) | |
| 8-11 | 96 | 7 (7.3%) | 0.40 (0.08-2.07) | |
| 12-16 | 78 | 3 (3.8%) | 0.34 (0.05-2.27) | |
| 17+ | 83 | 2 (2.4%) | 1.52 (0.32-7.21) | |
| **In the past week, left home to [b]:** | | | | |
| **Work** | 187 | 11 (5.9%) | 2.51 (0.81-7.73) | 0.11 |
| **Do groceries** | 364 | 17 (4.7%) | 0.22 (0.05-1.00) | 0.049 |
| **Visit family or friends** | 190 | 11 (5.8%) | 2.53 (0.86-7.43) | 0.092 |
| **Walk the dog or go outside with kids** | 58 | 2 (3.4%) | 0.68 (0.13-3.55) | 0.64 |
| **Walk or exercise outside** | 234 | 6 (2.6%) | 0.08 (0.03-0.26) | <0.001 |
| **Take care of someone** | 51 | 2 (3.9%) | 0.35 (0.07-1.74) | 0.20 |
| **Pick up prescription medicines or visit** | 97 | 7 (7.2%) | 0.90 (0.28-2.95) | 0.86 |
| **Attend religious services** | 13 | 2 (15.4%) | 1.26 (0.23-6.86) | 0.79 |
| **Visit cultural place** | 16 | 1 (6.3%) | 0.29 (0.03-2.49) | 0.26 |
| **Visit bar or restaurant** | 88 | 3 (3.4%) | 0.17 (0.05-0.67) | 0.011 |
| **Indoor sports** | 51 | 2 (3.9%) | 0.75 (0.15-3.72) | 0.73 |
| **Visit recreational park** | 67 | 3 (4.5%) | 0.65 (0.12-3.47) | 0.61 |
| **Frequency of using public transportation in the past week [b]** | | | | 0.13 |
| 0 days | 211 | 12 (5.7%) | 1 | |
| 1-2 days | 111 | 5 (4.5%) | 0.29 (0.08-1.04) | |
| 3-4 days | 45 | 3 (6.7%) | 0.44 (0.10-1.98) | |
| 5-7 days | 32 | 2 (6.3%) | 1.88 (0.27-13.04) | |

| | | | | |
|---|---|---|---|---|
| **Number of unique visitors at home in the past week** [b] | | | | 0.029 |
| 0 | 189 | 10 (5.3%) | 1 | |
| 1 | 81 | 3 (3.7%) | 0.54 (0.13-2.20) | |
| 2-4 | 97 | 6 (6.2%) | 4.68 (1.32-16.65) | |
| 5+ | 31 | 3 (9.7%) | 2.86 (0.54-15.15) | |
| **Travelled abroad in 2020** [b] | | | | 0.12 |
| No | 269 | 12 (4.5%) | 1 | |
| Yes | 129 | 10 (7.8%) | 2.76 (0.77-9.89) | |

**Abbreviations:** CI, confidence interval; HELIUS, Healthy Life in an Urban Setting; OR, odds ratio
[a] Those with an equivocal test result were excluded from this analysis [b] Measured at COVID-1 visit (2020) [c] Measured at baseline (2011-2015) [d] Quartiles [e] Presumed higher exposure categories had priority, i.e. if someone was working in a school and as a careworker, they were categorized as a health worker. Caretakers were not included as a category because many had other jobs.

In multivariable analysis, the distribution of migration generation was skewed to mostly one group and hence were not included. The ORs for having a household member suspected of infection, walk or exercise outside, living with a child 4-12 years old, leaving home to visit bar or restaurant, and household size were extremely high with overinflated 95%CI, and hence were not included. The following variables were removed as they were no longer significant in the multivariable model: leaving home to work (*P*=0.97), traveling with public transport (0.71), leaving home to care for someone (0.63), visiting friends or family (0.53), occupational level (0.28), travelling abroad (0.22), leaving home to do groceries (0.14), labor participation (0.091), age (0.058), living with a child 13-17 years old (0.054).

**Table S5. Univariable analysis of potential determinants of SARS-CoV-2 seropositivity in Ghanaian participants, Amsterdam, the Netherlands, 24 June - 9 October 2020**

| Characteristic | Number per category | Number with antibodies (%) | OR (95% CI)[a] | *P*-value |
|---|---|---|---|---|
| **Sex** | | | | 0.46 |
| Male | 143 | 40 (28.0%) | 1 | |
| Female | 184 | 55 (29.9%) | 1.25 (0.69-2.29) | |
| **Per year increase in age in years on 1 January 2020**[c] | | | 1.02 (0.99-1.05) | 0.12 |
| **Migration generation** | | | | |
| 1st | 321 | 95 (29.6%) | Omitted | |
| 2nd | 6 | 0 (0%) | Omitted | |
| **Month of study visit**[b] | | | | 0.026 |
| June | 0 | | Omitted | |
| July | 38 | 6 (15.8%) | 1 | |
| August | 133 | 47 (35.3%) | 4.13 (1.43-11.9) | |
| September | 123 | 35 (28.5%) | 2.06 (0.72-5.92) | |
| October | 33 | 7 (21.2%) | 1.57 (0.41-5.99) | |
| **City district**[c] | 38 | 6 (15.8%) | | 0.10 |
| Centre | 5 | 0 (0%) | Omitted | |
| East | 25 | 4 (16.0%) | 1.02 (0.19-5.41) | |
| West | 19 | 4 (21.1%) | 3.74 (0.52-26.92) | |
| South | 8 | 2 (25.0%) | 1.49 (0.27-8.38) | |
| New-West | 17 | 4 (23.5%) | 3.33 (1-11.11) | |
| Southeast | 251 | 81 (32.3%) | 1.02 (0.19-5.41) | |
| Other | 0 | | Omitted | |
| **Has obesity (BMI≥30.0)**[c] | | | | 0.77 |
| No | 225 | 66 (29.3%) | 1 | |
| Yes | 97 | 28 (28.9%) | 0.90 (0.45-1.81) | |
| **Educational level**[c] | | | | 0.23 |
| No school/elementary school | 78 | 26 (33.3%) | 1 | |
| Lower vocational/ lower secondary school | 127 | 36 (28.3%) | 0.70 (0.33-1.50) | |
| Intermediary vocational/ intermediary secondary school | 72 | 20 (27.8%) | 0.39 (0.18-0.86) | |
| Higher vocational/university | 26 | 7 (26.9%) | 0.75 (0.23-2.47) | |
| Missing | 24 | 6 (25.0%) | | |
| **Labor participation**[c] | | | | 0.82 |
| Employed | 202 | 60 (29.7%) | 1 | |
| Not in workforce | 10 | 2 (20.0%) | 0.51 (0.09-3.08) | |
| Unemployed/on benefits | 59 | 17 (28.8%) | 1.34 (0.61-2.95) | |
| Disabled | 28 | 8 (28.6%) | 0.8 (0.32-2.01) | |
| Unknown/missing | 28 | 8 (28.6%) | 1.01 (0.39-2.58) | |
| **Elementary occupation**[c] | | | | 0.33 |
| No | 107 | 36 (33.6%) | 1 | |
| Yes | 162 | 43 (26.5%) | 1.29 (0.68-2.44) | |
| Missing | 58 | 16 (27.6%) | 0.75 (0.31-1.81) | |
| **Difficulty with Dutch language**[c] | | | | 0.010 |
| No | 40 | 9 (22.5%) | 1 | |
| Yes | 263 | 80 (30.4%) | 3.21 (1.32-7.78) | |
| **Health literacy (SBSQ)**[c] | | | | 0.74 |
| Adequate | 207 | 59 (28.5%) | 1 | |
| Low | 97 | 30 (30.9%) | 1.12 (0.58-2.15) | |

| | | | | |
|---|---|---|---|---|
| **Job setting[b,e]** | | | | 0.85 |
| No job / caretaker only | 89 | 20 (22.5%) | 1 | |
| Job with no contact within 1.5 meter | 65 | 19 (29.2%) | 1.66 (0.69-3.99) | |
| Other job with contact within 1.5 meter | 114 | 36 (31.6%) | 1.56 (0.71-3.43) | |
| Child care/schools/higher education | 10 | 3 (30.0%) | 1.93 (0.25-15.1) | |
| Bar/restaurant | 23 | 7 (30.4%) | 1.49 (0.44-4.96) | |
| Hospital/long-term care facility/Care worker elsewhere | 25 | 9 (36.0%) | 1.11 (0.37-3.28) | |
| **Caretaker [b]** | | | | 0.71 |
| No | 307 | 89 (29.0%) | 1 | |
| Yes | 19 | 5 (26.3%) | 0.80 (0.25-2.59) | |
| **Number of people in household [c]** | | | | 0.06 |
| 1 (Lives alone) | 46 | 6 (13%) | 1 | |
| 2 | 61 | 18 (30%) | 1.85 (0.63-5.50) | |
| 3 | 69 | 19 (28%) | 1.88 (0.62-5.70) | |
| 4 | 69 | 23 (33%) | 2.86 (0.96-8.48) | |
| ≥5 | 55 | 21 (38%) | 5.02 (1.59-15.86) | |
| **Lives with other people [b]** | 268 | 77 (28.7%) | 0.94 (0.47-1.91) | 0.87 |
| **Partner** | 124 | 38 (30.6%) | 1.28 (0.68-2.39) | 0.45 |
| **Children up to 3 years old** | 26 | 11 (42.3%) | 2.54 (1.00-6.46) | 0.050 |
| **Children 4 through 12 years old** | 81 | 25 (30.9%) | 1.17 (0.59-2.33) | 0.65 |
| **Children 13 through 17 years old** | 76 | 28 (36.8%) | 1.97 (1.02-3.80) | 0.045 |
| **Children 18+ years old** | 114 | 37 (32.5%) | 1.52 (0.84-2.73) | 0.16 |
| **Parents or parents-in-law** | 8 | 0 (0%) | Omitted | |
| **Other adults** | 58 | 19 (32.8%) | 1.08 (0.51-2.30) | 0.83 |
| **Household member/steady partner with suspected infection [b]** | | | | 0.79 |
| N.A./No | 311 | 75 (28.0%) | 1 | |
| Yes | 15 | 4 (26.7%) | 1.20 (0.30-4.78) | |
| **Number of times left home in the past week [b,d]** | | | | 0.87 |
| 0-7 | 120 | 34 (28.3%) | 1 | |
| 8-11 | 118 | 33 (28.0%) | 0.90 (0.45-1.77) | |
| 12-16 | 58 | 16 (27.6%) | 1.07 (0.43-2.63) | |
| 17+ | 30 | 11 (36.7%) | 0.67 (0.24-1.89) | |
| **In the past week, left home to [b]:** | | | | |
| **Work** | 192 | 66 (34.4%) | 1.91 (1.01-3.60) | 0.045 |
| **Do groceries** | 294 | 84 (28.6%) | 1.29 (0.54-3.09) | 0.56 |
| **Visit family or friends** | 81 | 21 (25.9%) | 0.40 (0.21-0.78) | 0.007 |
| **Walk the dog or go outside with kids** | 22 | 10 (45.5%) | 2.27 (0.87-5.95) | 0.09 |
| **Walk or exercise outside** | 207 | 58 (28.0%) | 0.75 (0.40-1.40) | 0.37 |
| **Take care of someone** | 14 | 4 (28.6%) | 1.22 (0.36-4.08) | 0.75 |
| **Pick up prescription medicines or visit** | 70 | 20 (28.6%) | 0.82 (0.39-1.74) | 0.61 |
| **Attend religious service** | 128 | 49 (38.3%) | 2.76 (1.49-5.11) | 0.001 |
| **Visit cultural place** | 3 | 1 (33.3%) | 0.51 (0.04-5.91) | 0.59 |
| **Visit bar or restaurant** | 19 | 4 (21.1%) | 0.35 (0.11-1.15) | 0.082 |
| **Indoor sports** | 27 | 10 (37.0%) | 0.79 (0.30-2.10) | 0.63 |
| **Visit recreational park** | 17 | 4 (23.5%) | 0.79 (0.14-4.45) | 0.79 |
| **Frequency of using public transportation in the past week [b]** | | | | 0.90 |
| 0 days | 116 | 31 (26.7%) | 1 | |
| 1-2 days | 73 | 20 (27.4%) | 0.73 (0.31-1.72) | |
| 3-4 days | 38 | 13 (34.2%) | 1.01 (0.40-2.56) | |
| 5-7 days | 98 | 30 (30.6%) | 0.91 (0.43-1.93) | |

| | | | | |
|---|---|---|---|---|
| **Number of unique visitors at home in the past week** [b] | | | | 0.29 |
| 0 | 238 | 71 (29.8%) | 1 | |
| 1 | 41 | 7 (17.1%) | 0.47 (0.19-1.20) | |
| 2-4 | 40 | 14 (35.0%) | 0.57 (0.25-1.29) | |
| 5+ | 6 | 2 (33.3%) | 1.02 (0.17-5.93) | |
| **Travelled abroad in 2020** [b] | | | | 0.020 |
| No | 252 | 76 (30.2%) | 1 | |
| Yes | 72 | 18 (25.0%) | 0.44 (0.22-0.88) | |

**Abbreviations:** CI, confidence interval; HELIUS, Healthy Life in an Urban Setting; OR, odds ratio
[a] Those with an equivocal test result were excluded from this analysis [b] Measured at COVID-1 visit (2020) [c] Measured at baseline (2011-2015) [d] Quartiles [e] Presumed higher exposure categories had priority, i.e. if someone was working in a school and as a careworker, they were categorized as a health worker. Caretakers were not included as a category because many had other jobs.

In multivariable analysis, the following variables were removed as they were no longer significant in the multivariable model: living with a child 18+ years old (*P*=0.92), leaving home to visit bar or restaurant (0.91), travelling abroad (0.66), living with a child 13-17 years old (0.51), visiting friends or family (0.22), walk the dog or go outside with kids (0.15), difficulty with Dutch language (0.11), district (0.09).

**Table S6. Univariable analysis of potential determinants of SARS-CoV-2 seropositivity in Turkish participants, Amsterdam, the Netherlands, 24 June - 9 October 2020**

| Characteristic | Number per category | Number with antibodies (%) | OR (95% CI)[a] | *P*-value |
|---|---|---|---|---|
| **Sex** | | | | 0.63 |
| Male | 183 | 14 (7.7%) | 1 | |
| Female | 225 | 16 (7.1%) | 1.23 (0.53-2.90) | |
| **Per year increase in age in years on 1 January 2020**[c] | | | 0.97 (0.93-1.01) | 0.15 |
| **Migration generation** | | | | 0.24 |
| 1st | 306 | 20 (6.5%) | 1 | |
| 2nd | 102 | 10 (9.8%) | 1.67 (0.71-3.89) | |
| **Month of study visit**[b] | | | | 0.40 |
| June | 0 | | Omitted | |
| July | 151 | 9 (6.0%) | 1 | |
| August | 88 | 5 (5.7%) | 0.38 (0.10-1.45) | |
| September | 126 | 13 (10.3%) | 1.11 (0.41-2.99) | |
| October | 43 | 3 (7.0%) | 1.18 (0.27-5.13) | |
| **City district**[c] | | | | 0.056 |
| Centre | 3 | 0 (0%) | 1 | |
| East | 66 | 10 (15.2%) | 0.87 (0.27-2.75) | |
| West | 66 | 8 (12.1%) | 0.40 (0.06-2.79) | |
| South | 30 | 2 (6.7%) | 0.23 (0.08-0.67) | |
| New-West | 231 | 9 (3.9%) | 1.22 (0.12-12.99) | |
| Southeast | 6 | 1 (16.7%) | 0.87 (0.27-2.75) | |
| Other | 0 | | Omitted | |
| **Has obesity (BMI≥30.0)**[c] | | | | 0.41 |
| No | 288 | 22 (7.6%) | 1 | |
| Yes | 114 | 7 (6.1%) | 1.5 (0.58-3.92) | |
| **Educational level**[c] | | | | 0.95 |
| No school/elementary school | 78 | 6 (7.7%) | 1 | |
| Lower vocational/ lower secondary school | 84 | 8 (9.5%) | 1.41 (0.41-4.85) | |
| Intermediary vocational/ intermediary secondary school | 124 | 9 (7.3%) | 1.17 (0.36-3.83) | |
| Higher vocational/university | 107 | 7 (6.5%) | 1.39 (0.38-5.06) | |
| Missing | 15 | 0 (0%) | Omitted | |
| **Labor participation**[c] | | | | 0.76 |
| Employed | 246 | 17 (6.9%) | 1 | |
| Not in workforce | 59 | 8 (13.6%) | 1.68 (0.63-4.46) | |
| Unemployed/on benefits | 62 | 4 (6.5%) | 1.23 (0.37-4.09) | |
| Disabled | 27 | 1 (3.7%) | 0.82 (0.10-6.71) | |
| Unknown/missing | 14 | 0 (0%) | Omitted | |
| **Elementary occupation**[c] | | | | 0.47 |
| No | 272 | 17 (6.3%) | 1 | |
| Yes | 52 | 4 (7.7%) | 2.08 (0.61-7.15) | |
| Missing | 84 | 9 (10.7%) | 1.41 (0.54-3.66) | |
| **Difficulty with Dutch language**[c] | | | | 0.96 |
| No | 188 | 12 (6.4%) | 1 | |
| Yes | 206 | 17 (8.3%) | 1.02 (0.42-2.46) | |
| **Health literacy (SBSQ)**[c] | | | | 0.88 |
| Adequate | 309 | 21 (6.8%) | 1 | |
| Low | 87 | 9 (10.3%) | 1.07 (0.43-2.66) | |

| | | | | |
|---|---|---|---|---|
| **Job setting[b,e]** | | | | 1.00 |
| No job / caretaker only | 138 | 9 (6.5%) | 1 | |
| Job with no contact within 1.5 meter | 67 | 5 (7.5%) | 1.01 (0.27-3.69) | |
| Other job with contact within 1.5 meter | 129 | 8 (6.2%) | 0.87 (0.30-2.57) | |
| Child care/schools/higher education | 25 | 2 (8.0%) | 1.02 (0.14-7.45) | |
| Bar/restaurant | 6 | 1 (16.7%) | 0.99 (0.10-10.17) | |
| Hospital/long-term care facility/Care worker elsewhere | 41 | 5 (12.2%) | 1.18 (0.32-4.38) | |
| **Caretaker [b]** | | | | 0.076 |
| No | 362 | 23 (6.4%) | 1 | |
| Yes | 44 | 7 (15.9%) | 2.63 (0.9-7.67) | |
| **Number of people in household [c]** | | | | 0.43 |
| 1 (Lives alone) | 53 | 2 (3.8%) | 1 | |
| 2 | 67 | 2 (3.0%) | 1.45 (0.19-11.11) | |
| 3 | 81 | 5 (6.2%) | 2.17 (0.37-12.65) | |
| 4 | 113 | 10 (8.8%) | 2.71 (0.53-13.80) | |
| ≥5 | 81 | 10 (12.3%) | 4.11 (0.82-20.64) | |
| **Lives with other people [b]** | 345 | 27 (7.8%) | 1.00 (0.26-3.77) | 1.00 |
| **Partner** | 269 | 21 (7.8%) | 0.98 (0.40-2.38) | 0.96 |
| **Children up to 3 years old** | 38 | 3 (7.9%) | 1.46 (0.40-5.32) | 0.56 |
| **Children 4 through 12 years old** | 84 | 6 (7.1%) | 0.67 (0.23-2.00) | 0.48 |
| **Children 13 through 17 years old** | 76 | 6 (7.9%) | 0.92 (0.32-2.66) | 0.88 |
| **Children 18+ years old** | 172 | 14 (8.1%) | 1.29 (0.55-3.00) | 0.56 |
| **Parents or parents-in-law** | 31 | 3 (9.7%) | 0.76 (0.21-2.81) | 0.68 |
| **Other adults** | 18 | 2 (11.1%) | 1.58 (0.32-7.89) | 0.58 |
| **Household member/steady partner with suspected infection [b]** | | | | <0.001 |
| N.A./No | 359 | 16 (4.5%) | 1 | |
| Yes | 46 | 14 (30.4%) | 9.15 (3.7-22.63) | |
| **Number of times left home in the past week [b,d]** | | | | 0.73 |
| 0-7 | 106 | 6 (5.7%) | 1 | |
| 8-11 | 97 | 10 (10.3%) | 1.69 (0.51-5.57) | |
| 12-16 | 103 | 7 (6.8%) | 0.92 (0.25-3.40) | |
| 17+ | 100 | 7 (7.0%) | 1.20 (0.33-4.37) | |
| **In the past week, left home to [b]:** | | | | |
| **Work** | 190 | 19 (10.0%) | 1.59 (0.66-3.83) | 0.30 |
| **Do groceries** | 360 | 27 (7.5%) | 2.21 (0.56-8.73) | 0.26 |
| **Visit family or friends** | 215 | 17 (7.9%) | 1.14 (0.48-2.67) | 0.77 |
| **Walk the dog or go outside with kids** | 79 | 5 (6.3%) | 0.41 (0.13-1.27) | 0.12 |
| **Walk or exercise outside** | 264 | 23 (8.7%) | 3.53 (1.41-8.83) | 0.007 |
| **Take care of someone** | 37 | 5 (13.5%) | 2.07 (0.66-6.46) | 0.21 |
| **Pick up prescription medicines or visit** | 84 | 8 (9.5%) | 1.38 (0.53-3.61) | 0.51 |
| **Attend religious service** | 62 | 5 (8.1%) | 0.73 (0.26-2.10) | 0.56 |
| **Visit cultural place** | 14 | 3 (21.4%) | 3.81 (0.77-18.83) | 0.10 |
| **Visit bar or restaurant** | 127 | 9 (7.1%) | 0.76 (0.29-2.02) | 0.58 |
| **Indoor sports** | 42 | 6 (14.3%) | 1.81 (0.58-5.67) | 0.31 |
| **Visit recreational park** | 81 | 7 (8.6%) | 0.97 (0.35-2.68) | 0.95 |
| **Frequency of using public transportation in the past week [b]** | | | | 0.13 |
| 0 days | 307 | 19 (6.2%) | 1 | |
| 1-2 days | 72 | 7 (9.7%) | 2.76 (1.01-7.51) | |
| 3-4 days | 14 | 2 (14.3%) | 3.35 (0.64-17.39) | |
| 5-7 days | 13 | 2 (15.4%) | 2.72 (0.47-15.72) | |

| | | | | |
|---|---|---|---|---|
| **Number of unique visitors at home in the past week** [b] | | | | 0.055 |
| 0 | 207 | 18 (8.7%) | 1 | |
| 1 | 48 | 1 (2.1%) | 0.09 (0.01-0.69) | |
| 2-4 | 109 | 8 (7.3%) | 0.96 (0.34-2.68) | |
| 5+ | 41 | 3 (7.3%) | 0.33 (0.09-1.25) | |
| **Travelled abroad in 2020** [b] | | | | 0.73 |
| No | 234 | 17 (7.3%) | 1 | |
| Yes | 172 | 13 (7.6%) | 1.17 (0.49-2.78) | |

**Abbreviations:** CI, confidence interval; HELIUS, Healthy Life in an Urban Setting; OR, odds ratio
[a] Those with an equivocal test result were excluded from this analysis [b] Measured at COVID-1 visit (2020) [c] Measured at baseline (2011-2015) [d] Quartiles [e] Presumed higher exposure categories had priority, i.e. if someone was working in a school and as a careworker, they were categorized as a health worker. Caretakers were not included as a category because many had other jobs.

In multivariable analysis, the following variables were removed as they were no longer significant in the multivariable model: visit cultural place (P=0.89), walk the dog or go outside with kids (0.38), being a caretaker (0.27), number of unique visitors past week (0.26), age (0.20), household size (0.12), district (0.11)

**Table S7. Univariable analysis of potential determinants of SARS-CoV-2 seropositivity in Moroccan participants, Amsterdam, the Netherlands, 24 June - 9 October 2020**

| Characteristic | Number per category | Number with antibodies (%) | OR (95% CI)[a] | *P*-value |
|---|---|---|---|---|
| **Sex** | | | | 0.093 |
| Male | 172 | 11 (6.4%) | 1 | |
| Female | 219 | 21 (9.6%) | 2.26 (0.87-5.86) | |
| **Per year increase in age in years on 1 January 2020**[c] | | | 1.00 (0.96-1.03) | 0.90 |
| **Migration generation** | | | | 0.23 |
| 1st | 297 | 23 (7.7%) | 1 | |
| 2nd | 94 | 9 (9.6%) | 1.74 (0.71-4.25) | |
| **Month of study visit**[b] | | | | 0.24 |
| June | 32 | 4 (12.5%) | 1 | |
| July | 168 | 7 (4.2%) | 0.41 (0.09-1.79) | |
| August | 85 | 7 (8.2%) | 0.72 (0.17-3.06) | |
| September | 74 | 12 (16.2%) | 1.66 (0.4-6.85) | |
| October | 32 | 2 (6.3%) | 0.78 (0.12-5.00) | |
| **City district** [c] | | | | 0.040 |
| Centre | 12 | 2 (16.7%) | 1 | |
| East | 94 | 10 (10.6%) | 0.83 (0.15-4.76) | |
| West | 82 | 10 (12.2%) | 1.77 (0.30-10.35) | |
| South | 38 | 2 (5.3%) | 0.32 (0.03-2.98) | |
| New-West | 145 | 8 (5.5%) | 0.34 (0.06-1.99) | |
| Southeast | 19 | 0 (0%) | Omitted | |
| Other | 1 | 0 (0%) | Omitted | |
| **Has obesity (BMI≥30.0)** [c] | | | | 0.96 |
| No | 288 | 25 (8.7%) | | |
| Yes | 96 | 7 (7.3%) | 1.03 (0.37-2.90) | |
| **Educational level** [c] | | | | 0.072 |
| No school/elementary school | 89 | 6 (6.7%) | 1 | |
| Lower vocational/ lower secondary school | 64 | 4 (6.3%) | 1.30 (0.34-5.00) | |
| Intermediary vocational/ intermediary secondary school | 123 | 11 (8.9%) | 1.47 (0.48-4.47) | |
| Higher vocational/university | 94 | 7 (7.4%) | 1.39 (0.41-4.72) | |
| Missing | 21 | 4 (19.0%) | 8.52 (1.92-37.78) | |
| **Labor participation** [c] | | | | 0.011 |
| Employed | 227 | 16 (7.0%) | 1 | |
| Not in workforce | 63 | 5 (7.9%) | 1.48 (0.49-4.47) | |
| Unemployed/on benefits | 56 | 4 (7.1%) | 1.03 (0.3-3.48) | |
| Disabled | 22 | 2 (9.1%) | 1.01 (0.19-5.5) | |
| Unknown/missing | | | 9.2 (2.68-31.54) | |
| **Elementary occupation** [c] | | | | 0.003 |
| No | 259 | 16 (6.2%) | 1 | |
| Yes | 46 | 4 (8.7%) | 1.49 (0.45-4.99) | |
| Missing | 86 | 12 (14.0%) | 4.69 (1.93-11.43) | |
| **Difficulty with Dutch language** [c] | | | | 0.33 |
| No | 210 | 14 (6.7%) | 1 | |
| Yes | 160 | 14 (8.8%) | 1.53 (0.65-3.62) | |
| **Health literacy (SBSQ)** [c] | | | | 0.50 |
| Adequate | 305 | 21 (6.9%) | 1 | |
| Low | 64 | 7 (10.9%) | 1.39 (0.54-3.58) | |

| | | | | |
|---|---|---|---|---|
| **Job setting**[b,e] | | | | 0.017 |
| No job / caretaker only | 131 | 12 (9.2%) | 1 | |
| Job with no contact within 1.5 meter | 54 | 7 (13.0%) | 0.82 (0.27-2.47) | |
| Other job with contact within 1.5 meter | 113 | 3 (2.7%) | 0.15 (0.04-0.64) | |
| Child care/schools/higher education | 48 | 8 (16.7%) | 2.15 (0.68-6.80) | |
| Bar/restaurant | 7 | 1 (14.3%) | 0.88 (0.09-8.16) | |
| Hospital/long-term care facility/Care worker elsewhere | 35 | 1 (2.9%) | 0.13 (0.02-1.08) | |
| **Caretaker** [b] | | | | 0.42 |
| No | 335 | 26 (7.8%) | 1 | |
| Yes | 53 | 6 (11.3%) | 1.59 (0.51-4.90) | |
| **Number of people in household** [c] | | | | 0.41 |
| 1 (Lives alone) | 52 | 4 (7.7%) | 1 | |
| 2 | 64 | 1 (1.6%) | 0.24 (0.03-2.26) | |
| 3 | 40 | 3 (7.5%) | 0.56 (0.11-2.76) | |
| 4 | 71 | 8 (11.3%) | 1.47 (0.39-5.51) | |
| ≥5 | 142 | 12 (8.5%) | 1.20 (0.34-4.17) | |
| **Lives with other people** [b] | 318 | 27 (8.5%) | 1.09 (0.32-3.72) | 0.90 |
| **Partner** | 256 | 22 (8.6%) | 0.85 (0.34-2.12) | 0.72 |
| **Children up to 3 years old** | 48 | 5 (10.4%) | 1.19 (0.41-3.45) | 0.74 |
| **Children 4 through 12 years old** | 104 | 10 (9.6%) | 1.16 (0.47-2.84) | 0.75 |
| **Children 13 through 17 years old** | 114 | 11 (9.6%) | 1.06 (0.44-2.53) | 0.90 |
| **Children 18+ years old** | 149 | 15 (10.1%) | 0.94 (0.41-2.15) | 0.89 |
| **Parents or parents-in-law** | 21 | 2 (9.5%) | 1.76 (0.36-8.69) | 0.49 |
| **Other adults** | 22 | 3 (13.6%) | 2.99 (0.74-12.08) | 0.12 |
| **Household member/steady partner with suspected infection** [b] | | | | <0.001 |
| N.A./No | 336 | 18 (5.4%) | 1 | |
| Yes | 51 | 14 (27.5%) | 5.14 (2.06-12.82) | |
| **Number of times left home in the past week** [b,d] | | | | 0.020 |
| 0-7 | 99 | 15 (15.2%) | 1 | |
| 8-11 | 89 | 5 (5.6%) | 0.20 (0.06-0.65) | |
| 12-16 | 88 | 7 (8.0%) | 0.33 (0.11-0.97) | |
| 17+ | 113 | 5 (4.4%) | 0.28 (0.08-1.00) | |
| **In the past week, left home to** [b]**:** | | | | |
| **Work** | 171 | 9 (5.3%) | 0.46 (0.17-1.28) | 0.14 |
| **Do groceries** | 363 | 28 (7.7%) | 0.40 (0.11-1.37) | 0.14 |
| **Visit family or friends** | 226 | 15 (6.6%) | 0.50 (0.21-1.19) | 0.12 |
| **Walk the dog or go outside with kids** | 77 | 6 (7.8%) | 0.98 (0.31-3.13) | 0.97 |
| **Walk or exercise outside** | 268 | 19 (7.1%) | 1.02 (0.43-2.4) | 0.96 |
| **Take care of someone** | 57 | 5 (8.8%) | 0.90 (0.30-2.68) | 0.84 |
| **Pick up prescription medicines or visit** | 89 | 9 (10.1%) | 1.25 (0.44-3.56) | 0.68 |
| **Attend religious service** | 32 | 2 (6.3%) | 0.58 (0.12-2.82) | 0.50 |
| **Visit cultural place** | 15 | 0 (0%) | Omitted | |
| **Visit bar or restaurant** | 130 | 9 (6.9%) | 0.90 (0.33-2.44) | 0.84 |
| **Indoor sports** | 42 | 4 (9.5%) | 1.32 (0.29-6.10) | 0.72 |
| **Visit recreational park** | 94 | 5 (5.3%) | 0.55 (0.19-1.60) | 0.27 |
| **Frequency of using public transportation in the past week** [b] | | | | 0.18 |
| 0 days | 256 | 22 (8.6%) | 1 | |
| 1-2 days | 90 | 6 (6.7%) | 0.41 (0.15-1.13) | |
| 3-4 days | 25 | 3 (12.0%) | 0.58 (0.15-2.19) | |
| 5-7 days | 18 | 1 (5.6%) | 0.20 (0.03-1.66) | |

| | | | | |
|---|---|---|---|---|
| **Number of unique visitors at home in the past week** [b] | | | | 0.11 |
| 0 | 206 | 22 (10.7%) | 1 | |
| 1 | 45 | 1 (2.2%) | 0.13 (0.02-1.02) | |
| 2-4 | 102 | 7 (6.9%) | 1.08 (0.39-3.01) | |
| 5+ | 34 | 2 (5.9%) | 0.29 (0.06-1.35) | |
| **Travelled abroad in 2020** [b] | | | | 0.11 |
| No | 228 | 15 (6.6%) | 1 | |
| Yes | 160 | 16 (10.0%) | 1.99 (0.85-4.67) | |

**Abbreviations:** CI, confidence interval; HELIUS, Healthy Life in an Urban Setting; OR, odds ratio
[a] Those with an equivocal test result were excluded from this analysis [b] Measured at COVID-1 visit (2020) [c] Measured at baseline (2011-2015) [d] Quartiles [e] Presumed higher exposure categories had priority, i.e. if someone was working in a school and as a careworker, they were categorized as a health worker. Caretakers were not included as a category because many had other jobs.

In multivariable analysis, the distribution of district was skewed to mostly one group and hence were not included. The following variables were removed as they were no longer significant in the multivariable model: sex ($P$=0.93), living with other adults (0.83), number of unique visitors past week (0.82), leaving home to work (0.54), labor participation (0.57), visiting friends or family (0.87), education level (0.88), job setting (0.56), travelling abroad (0.28), groceries (0.30), number of time left house (0.12).
